# Escape from X-chromosome inactivation increases disease risk and drives sex-differential genetic effects: a population-scale PheXWAS

**DOI:** 10.64898/2026.11.20.26368570

**Authors:** Miguel Ángel Pérez-Elena, Cecilia Salvoro, Daniel Matías-Sánchez, Leila Satari, Montserrat Puiggròs, Daniel Zugasti-Fernandez, Lorena Alonso-Parrilla, David Torrents

## Abstract

X chromosome (Xchr) contribution to complex traits remains incomplete, due to its unique inheritance pattern and its inactivation mechanism. Therefore, we gathered data from 800,000 individuals from the UK Biobank and dbGaP, and performed a sex–stratified phenome–wide association study of Xchr variation across 3,741 complex traits. Heritability analyses revealed sex-specific patterns and an overall involvement of Xchr in complex phenotypes. We identified 6,531 loci significantly associated with 402 complex traits, with a higher proportion of associations consistent with dominant inheritance than observed for autosomes. Although effect directions were largely concordant between sexes, loci with significant effect-size differences were enriched for escape-gene associations. Escape genes were depleted among trait-associated loci and strongly enriched for risky disease effects. These association patterns enabled prediction of escape status for unannotated genes. This overview of Xchr contribution to complex traits suggests inactivation escape as a clear risky feature for females.

## Introduction

Commonly placed at the end of human karyotypes, the X chromosome (Xchr) is the eighth largest chromosome, accounting for nearly 5% of the genome, approximately 6% of all annotated genes, and about 3% of known human genetic variation^1,2^. Beyond its size and gene content, the distinctive biology of this sex chromosome, together with the effects of its allele-specific inactivation process, positions its genomic analysis as a promising avenue for advancing our understanding of the genetic and molecular mechanisms underlying complex disorders^3,4^.

Genome-wide association studies (GWAS) have identified thousands of variants associated with complex traits^5,6^. But one important gap of information affects our understanding about the role of Xchr in common diseases. Currently, only about 0.4% of all complex-trait-associated variants reported in the GWAS Catalog map to Xchr, a striking underrepresentation. This is nine times lower than expected when compared with autosomes of similar genetic variability, such as chromosome 16, which account for 4% of associations^7^. This underrepresentation could be due to the traditional exclusion of the Xchr from most GWASs because of the additional methodological costs^8,9^. The particular pattern of inheritance of this sexual chromosome and the effects of its specific allelic inactivation processes in females usually translate into allelic imbalances between sexes and the loss of statistical power during genetic association studies^6,10,11^. The presence of a single copy of this chromosome in males and, effectively, also in females through allelic inactivation, requires a different analysis strategy than that applied to autosomes. To address this complexity alternative protocols and analytical strategies have been proposed to investigate the contribution of Xchr across multiple complex traits^12–16^. These approaches commonly include sex-stratified analyses and sex-specific genotype-coding schemes that adapt the statistical model to the distinct allele composition in males and females^12–14^. Whether this marked underrepresentation reflects a genuinely smaller contribution of Xchr to complex trait biology or, instead, results from its historical exclusion from genetic studies remains unknown^8,9^.

Beyond the methodological challenges in analyzing the contribution of Xchr in complex diseases, understanding how escape from inactivation impacts complex disease susceptibility remains an unsolved challenge^3^. Although *XIST*-mediated inactivation of one chromosome balances gene dosage between sexes^17,18^, approximately 15-30% of X-linked genes escape this silencing^19–22^. Such escape is expected to generate sex-biased transcriptional profiles in different tissues and cell types^19,23,24^, that have been used to systematically classify X-linked genes according to their escape or inactive status^21,25,26^. Beyond dosage effects, escape from Xchr inactivation (XCI) has also been proposed as a contributor to sex-differential susceptibility in complex phenotypes such as cancer and autism^27,28^.

Despite multiple efforts to classify Xchr genes as subject to escaping or inactivation, the condition for more than 1,483 genes, according to GENECODE^29^, is inconclusive^19,26^. The creation of classification models were basing their decisions so far on multiple genomic elements surrounding the test gene, such as regulatory elements and repeats, which are correlated with escape events^30–32^. Apart from the assessment of expression and the genomic context, no other features, such as the relationship with disease, have been explored to achieve the classification of Xchr genes.

In this study, we aim to characterize the genetic contribution of Xchr to complex diseases and clinical traits, while bridging the existing gap in understanding how X-linked genetic architecture and X-inactivation influence disease risk. Leveraging UK Biobank and dbGaP resources for nearly 800 thousand individuals, we perform a sex-stratified phenome-wide association analysis of the non-PAR Xchr region across 3,741 complex traits. We quantify and characterize the contribution of Xchr to these diseases at the level of heritability and associated loci. Beyond this global characterization, we also investigate how X-inactivation and escape influence genetic association and risk, and test whether these disease-associated patterns can distinguish gene classes using a machine learning framework. The resulting insights refine our understanding of X–chromosome inheritance, clarify its role in complex disease risk, and open avenues for more precise sex–specific prevention and therapeutic strategies.

## Results

### Overall strategy

To study the role of the non-pseudoautosomal (non-PAR) region of Xchr in complex traits, we aggregated and harmonized genetic and phenotypic data from 792,968 individuals (428,493 females and 364,475 males; Supplemental Table 1) sourced from dbGaP and the UK Biobank (UKBK) and applied non-PAR Xchr-specific quality control^16^ (Methods). Leveraging five genetic reference panels^15,33^ (Methods), we imputed 418,231 as average across cohort and 469,009 specifically on the UKBK of high-quality Xchr variants (INFO≥0.7; Supplemental Table 2), comprising single-nucleotide variants, small insertions/deletions, and larger structural rearrangements^1^, which served as the basis for downstream genetic analyses (Figure 1). We curated the phenotypic data by filtering case and control subjects for binary disease traits. In total, 3,741 complex traits passed these filters, including 3,387 diseases and 354 quantitative disease-related clinical measures. From these, 63 and 24 are female and male-exclusive phenotypes, respectively (Supplemental Table 3). Using sex-specific X-chromosome genotype coding, we performed sex-stratified phenome-wide association studies (PheXWAS) with REGENIE^34^, evaluating additive, dominant, and recessive models in females, and a hemizygous model in males (Methods). To complement sex-stratified analysis, we also meta-analyzed the results. The resulting associations were then used to investigate the contribution of Xchr to complex phenotypes by comparing its heritability estimates with all chromosomes, and by identifying and characterizing specific X-linked locus-gene-phenotype associations.

**Figure 1.**
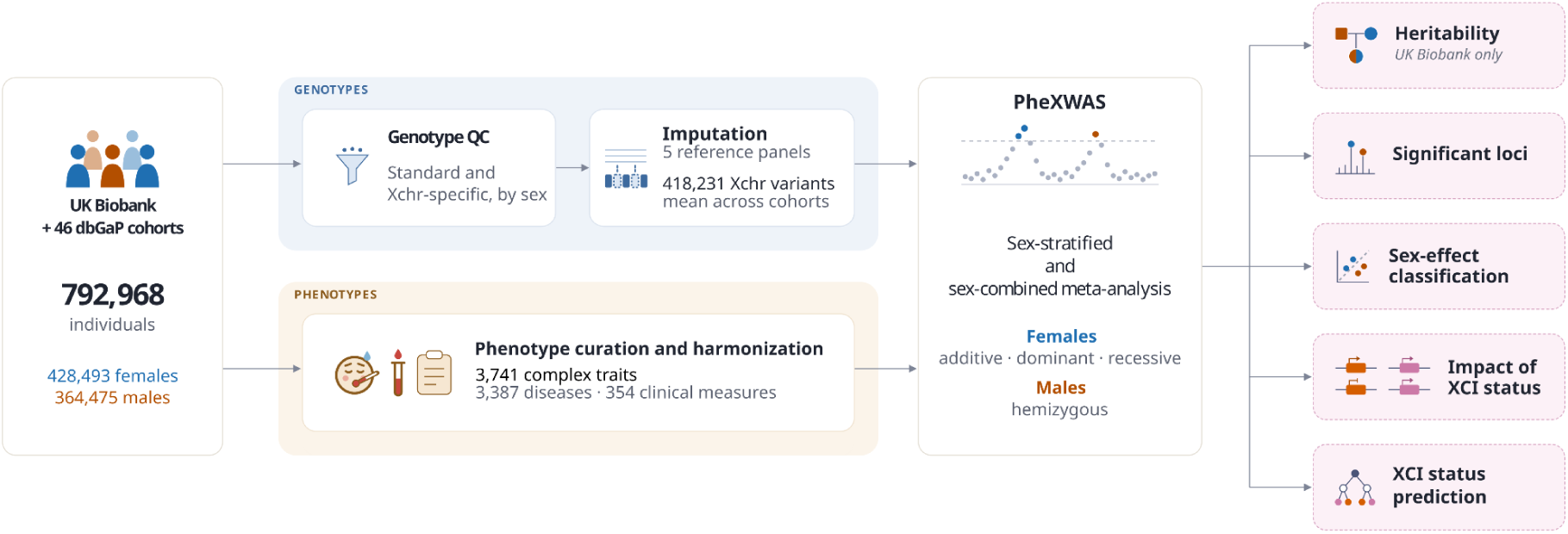
Overall strategy. We aggregated and harmonized genetic and phenotypic data from 792,968 individuals from 46 cohorts of dbGaP and the UK Biobank. Autosomal standard and Xchr-specific quality control were performed before the genotype imputation, which were applied to the filtered genotype array data using five genetic reference panels^15,16,33^ (Methods). An average of 418,231 high-quality variants across cohorts were tested for association with 3,741 complex traits. REGENIE^34^ was used to perform sex-stratified association studies, where females were evaluated under additive, dominant, and recessive inheritance models, and males under the hemizygous model. Sex-stratified results were meta-analyzed^50^. The resulting summary statistics were used to investigate the contribution of the Xchr to complex phenotypes. Right, downstream analyses: chromosome-stratified genome-wide (solid box; UK Biobank only) or restricted to the Xchr (dashed boxes), with the figures presenting each.

### Xchr contribution to complex traits

Within the UKBK cohort, we first calculated heritability metrics for all chromosomes separately for females (h^2^_F_) and males (h^2^_M_), across 3,663 complex traits with at least 5,000 individuals, using linkage disequilibrium score regression^35^ (LDSC; Methods). Overall, considering both sexes, we detected significant heritability (q ≤ 0.05) in at least one chromosome for 76% (2,780) of traits. On average, each chromosome contributed significantly to 37% of the traits, whereas Xchr showed a significant contribution (h²_X_) to 33% (929; Supplemental Table 4; Supplemental Figure 1). Estimates of h²_X_ varied substantially across traits, with an average of 0.002 and a range from 0.0003 to 0.0409. Beyond the overall h²_X_, sex-stratified h²_X_ distributions were comparable between females and males on their respective significant phenotypes (median_F_ = 0.0017; median_M_ = 0.0018; Mann–Whitney U *p* = 0.93; Supplemental Figure 2), although sex-specific patterns emerged across phenotype categories built on our data for better interpretability. In females, the highest median h²_X_ estimates were found for diseases classified under “Nervous system”, “Digestive system” and “Musculoskeletal/connective”. In contrast, in males, the highest median h²_X_ contributions were found within the “Nervous system”, “Digestive system” and “Mental disorders” (Figure 2A; Supplemental Figure 3). Across significantly associated phenotypes in both sexes, h²_X_ was consistently lower in males than females (median ratios male-to-female = 0.87; paired-samples t-test corrected *p* = 4.4×10⁻¹¹; Figure 2B; Supplemental Figure 4). In the disease category, 7 of 12 showed higher h²_X_ in females, with the largest difference in “Endocrine system” , whereas “Abnormal clinical findings” exhibited the largest difference in the opposite direction (Figure 2C).

**Figure 2.**
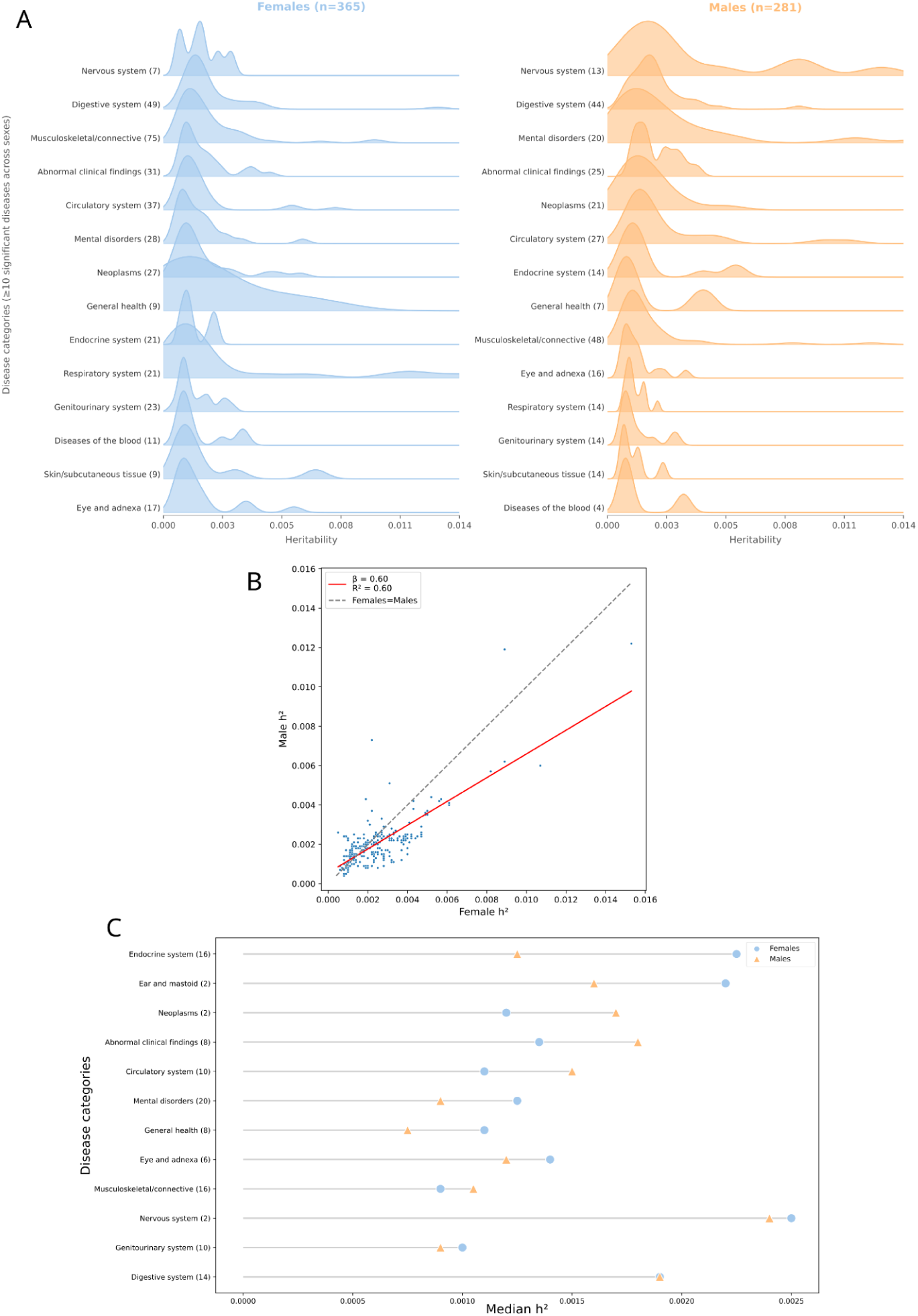
Chromosome X heritability estimates. **A)** Significant h²_X_ estimates across disease categories in females (left, n = 365 diseases) and males (right, n = 281), sorted by descending median h²_X_. Only categories with at least 10 significant diseases across both sexes are shown; numbers in parentheses give the number of diseases per category in each sex. **B)** Male versus female h²_X_ for the 259 phenotypes with significant h²_X_ in both sexes. The red line shows the linear regression (β = 0.60, R² = 0.60) and the dashed grey line the identity (equal h²_X_ in both sexes). **C)** Median h²_X_ per disease category in females (blue circles) and males (orange triangles); numbers in parentheses give the number of diseases per chapter.

To understand the contribution of Xchr relative to autosomes, we defined Chromosome Relative Involvement (CRI) as the percentage of total heritability (h²_T_) attributable to each chromosome, normalized by their corresponding number of imputed variants (Supplemental Table 5, Supplemental Figure 5). Within both sexes, the median CRI_X_ is significantly lower than the median CRI_A_, calculated from the distribution of per-autosome values (median CRI_X_ = 5.3%; median CRI_A_ = 8%; Mann–Whitney U *p* = 2.4×10⁻³¹; Supplemental Figure 6), ranking 6th in females and 2nd in males from the tail in contribution (Figure 3A). Furthermore, CRI_X_ values were approximately half of those derived from chromosomes with similar genetic variability, such as chromosome 16 (medians CRI_X_ = 5.3%; medians CRI_16_ = 11.9%; Brunner-Munzel test *p*_X,16_ = 2×10⁻⁴⁵).

**Figure 3.**
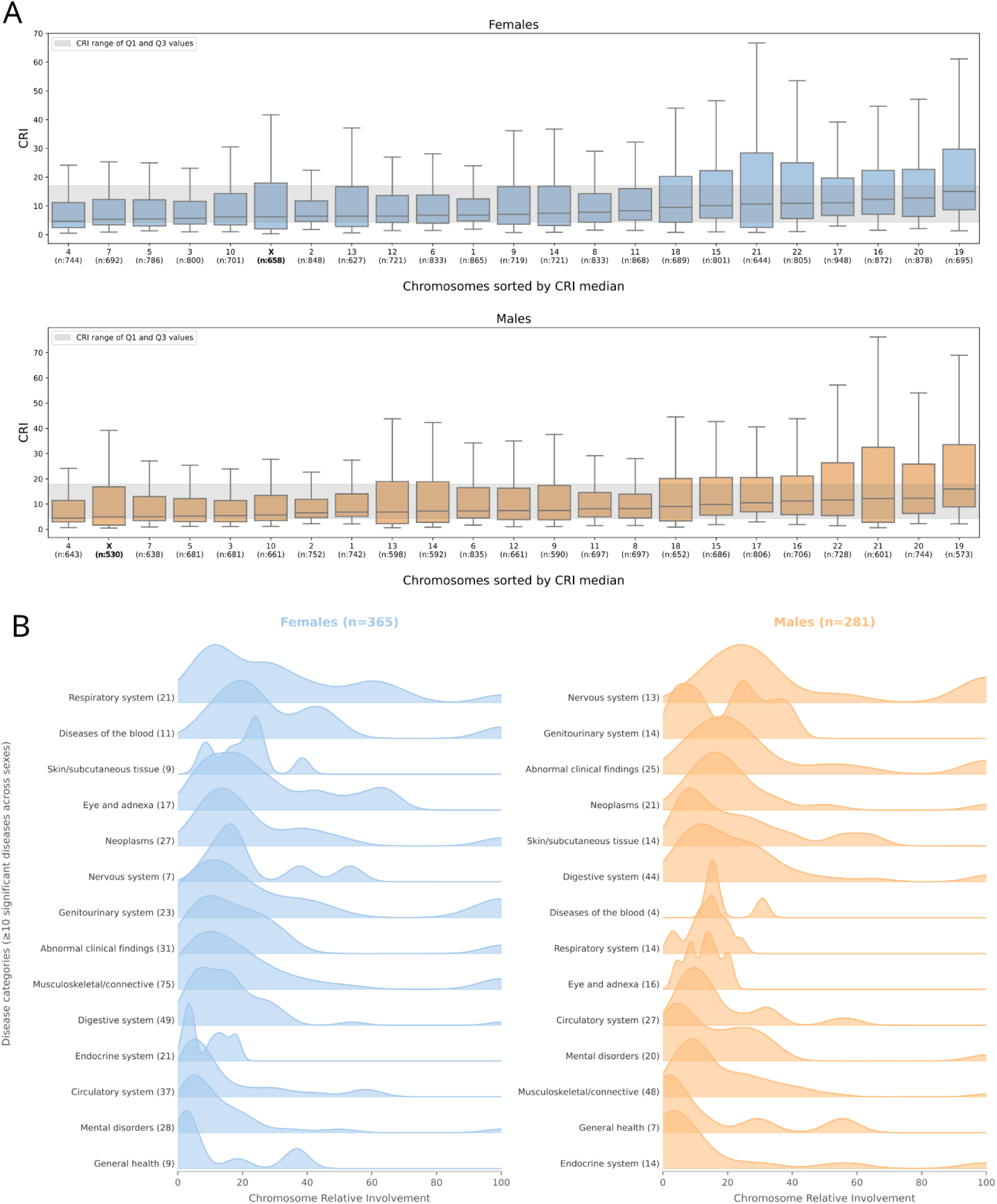
Chromosome Relative Involvement measures. **A)** CRI per chromosome across the 2,780 complex traits with significant heritability in at least one chromosome, in females (top) and males (bottom). Chromosomes are sorted by ascending median CRI and Xchr is shown in bold; n, number of traits with significant heritability for that chromosome. The grey band spans the first to third quartile across chromosomes. Outliers are not shown. **B)** Xchr CRI across disease categories in females (left, n = 365 diseases) and males (right, n = 281), sorted by descending median CRI.

Building on the comparison of X-chromosome contribution to that of the autosomes, we next examined how X-chromosome contribution to disease is distributed across disease categories. The highest median CRI_X_ in males was observed for “Nervous system” , “Genitourinary system”, and “Abnormal clinical findings”, whereas in females the highest values were observed for “Respiratory system” , “Diseases of the blood” and “Skin/subcutaneous tissue” (Figure 3B). Interestingly, 34 diseases exhibited heritability entirely attributable to Xchr (CRI_X_ = 100%), including “osteopathies in diseases classified elsewhere” (M90), “coagulation defects and inflammatory disease of the cervix uteri” (N72) in females; as well as “secondary malignant neoplasm of kidney and renal pelvis” (C79.0) and “Inflammatory polyneuropathy” (G61) in males (Supplemental Figure 7).

### Identification of X chromosome trait associated loci

Beyond global heritability measures, we next investigated the involvement of Xchr in complex traits at the level of significantly associated loci. These loci were extracted from the PheXWAS results across all tested complex traits using a *p*-value threshold derived from n-of-1 permutation test^36^ (Methods). For each female associated locus and disease, we selected the most likely associated inheritance model by prioritizing that with the lowest *p*-value (Figure 1). Under these criteria, we identify 205,638 significant variant–phenotype associations mapping to 6,531 unique independent loci, (r² > 0.8) across 402 complex traits (Figure 4A; Methods; Supplemental Table 6). A fraction of associations were discovered in coinciding phenotypes between UKBK and dbGaP, yielding a substantial cross–cohort replication (79% and 80%, respectively; Supplemental Table 7-8). Consistent with the allelic composition of the Xchr and the process of XCI in females, a substantial fraction (27%) of associations was best explained by the dominant inheritance model, showing approximately 1.6-fold more than previously observed for autosomes^15^. The recessive model explains 7% of the final results, 1.6-fold less than shown for autosomal variants (Figure 4B). In addition, we observed an important variation in the number of associated loci between phenotypes, with 21 traits accumulating more than 50 loci (Supplemental Figure 8). Among females, “mean platelet (thrombocyte) volume” and “menarche” showed the highest numbers of associated loci, with 233 and 129, respectively. Among males, “hair/balding pattern 4”, consistent with heritability results, and “testosterone levels” were associated with 297 and 128 loci, respectively (Figure 4A).

**Figure 4.**
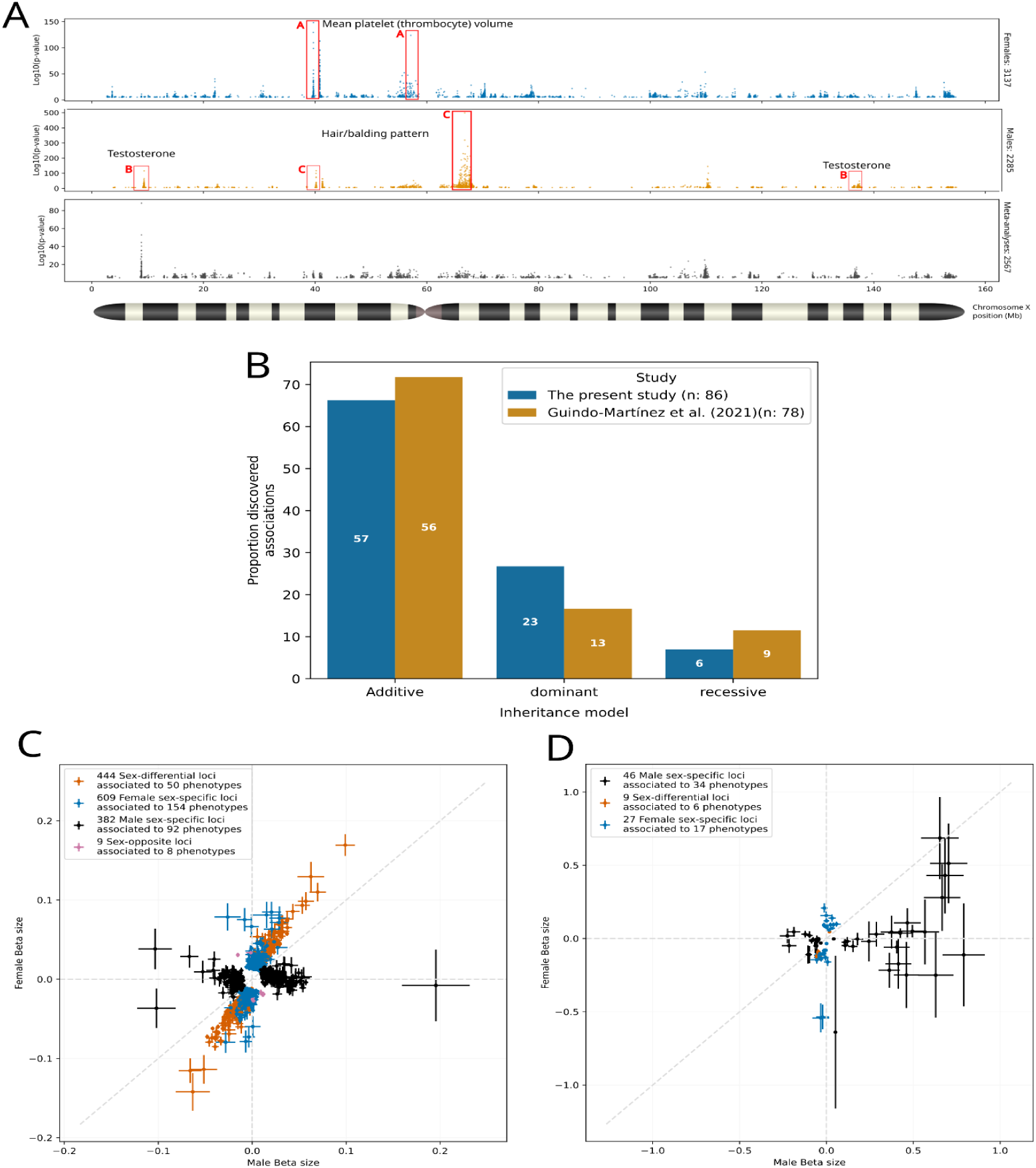
PheXWAS significant loci. **A)** Manhattan plots of significant associations lead variatns from the sex-stratified PheXWAS in females (top, blue; 3,137 loci), males (middle, orange; 2,285 loci), and from the meta-analysis (bottom, grey; 2,567 loci). Together they represent 6,531 unique independent loci (r² > 0.8) across 402 complex traits. Red boxes highlight examples of traits with more than 100 loci: A, “Mean platelet (thrombocyte) volume” in females; B, “Testosterone” and C, “hair/balding pattern” in males. **B)** Percentage of associations best explained by the additive, dominant and recessive models in this study (blue; n = 86 associations) and in Guindo-Martínez et al. (2021) (orange; n = 78 associations). Numbers inside the bars are counts. **C–D)** Female versus male effect sizes (β) of significant loci in non-sex-exclusive clinical measures (C) and diseases (D), coloured by sex-effect class: sex-differential (orange), female-specific (blue), male-specific (black) and sex-opposite (pink). Crosses show standard error of β; the dashed grey line shows equal effects in both sexes.

We next examined sex-specific patterns in greater detail for non-sex-exclusive traits^37^ (Methods). Significant associated loci were classified into five categories according to their patterns of association across sexes: sex-shared (no differences in association or effect), sex-differential (significantly different effect sizes in the same direction), sex-opposite (opposite directions of effect between sexes), female-specific (association detected only in females), and male-specific (association detected only in males). 39% of classifiable loci were shared between female and male, whereas the majority (61%) showed different behaviours between sexes. Specifically, 25.3% and 17% exhibited female- and male-specific association signals, respectively. Among the remaining loci, 18% showed significantly different effect sizes between sexes while retaining the same direction of effect. Only 9 loci (0.3%) with different effect sizes displayed opposite directions between sexes (Figure 4C-D; Supplemental Figure 9).

### Functional implications of Xchr–associated genes

To evaluate the functional landscape of Xchr-associated loci, we first mapped all 6,531 significant loci to 601 unique genes using colocalization analyses against cis-eQTL summaries statistics, complemented with gene proximity (Methods; Supplemental Table 9). These represent 28% of all genes in the Xchr, including coding, non-coding and microRNAs (Supplemental Table 10). A functional enrichment analysis of these trait-associated genes by considering both sexes together (Methods) shows 8 significant enriched functions (Supplemental Table 11), from which the top 3 correspond to “toll-like receptor 8 signaling pathway”, “neurexin family protein binding” and “Protein-protein interactions at synapses”. When stratifying the enrichment analyses by the genes mapped to sex-specific loci in the non-sex-exclusive phenotypes, we found only in males 35 significantly enriched pathways, including “cellular response to testosterone stimulus” and “Postsplicing complex” (Supplemental Table 12; Methods).

To evaluate whether Xchr preferably contributes to complex traits through specific tissues, we performed an enrichment analysis against expression profiles of 54 GTEx tissues (Methods; Supplemental Table 13). 27 tissues showed significant enrichment, with brain regions (e.g. hypothalamus, amygdala and caudate-basal ganglia) occupying the top four positions and accounting for 10 of the 27 (37%; Supplemental Figure 10). This pattern is consistent with the “Nervous system” phenotype category showing the largest h²_X_ contribution across trait categories (Supplemental Figure 3).

### Impact of chromosome X inactivation status in complex trait

To investigate the relationship between XCI status and phenotype susceptibility, we first characterized XCI patterns across all genes in Xchr identified as significantly associated with complex traits in females. Using multiple independent annotations^38^, we classified 563 genes into 275 inactive, 105 escapee, and 183 with uncertain XCI status (Supplemental Table 14, Methods). Genes with uncertain status were excluded from subsequent comparisons. We observed that escape genes accounted for only 25% of all loci with a trait association, compared with 39% of non-disease-related loci, indicating a significant underrepresentation of escapees among trait-association loci (χ² test *p* = 3×10⁻¹²⁷; Methods; Figure 5A). As expected the complementary trend was observed for inactive genes. Consistent with this depletion, the same was observed when using independent annotations from the GWAS Catalog with 29% of escapees mapping to trait-associated loci versus 39% mapping to non-disease loci (χ² test *p* = 0.02).

**Figure 5.**
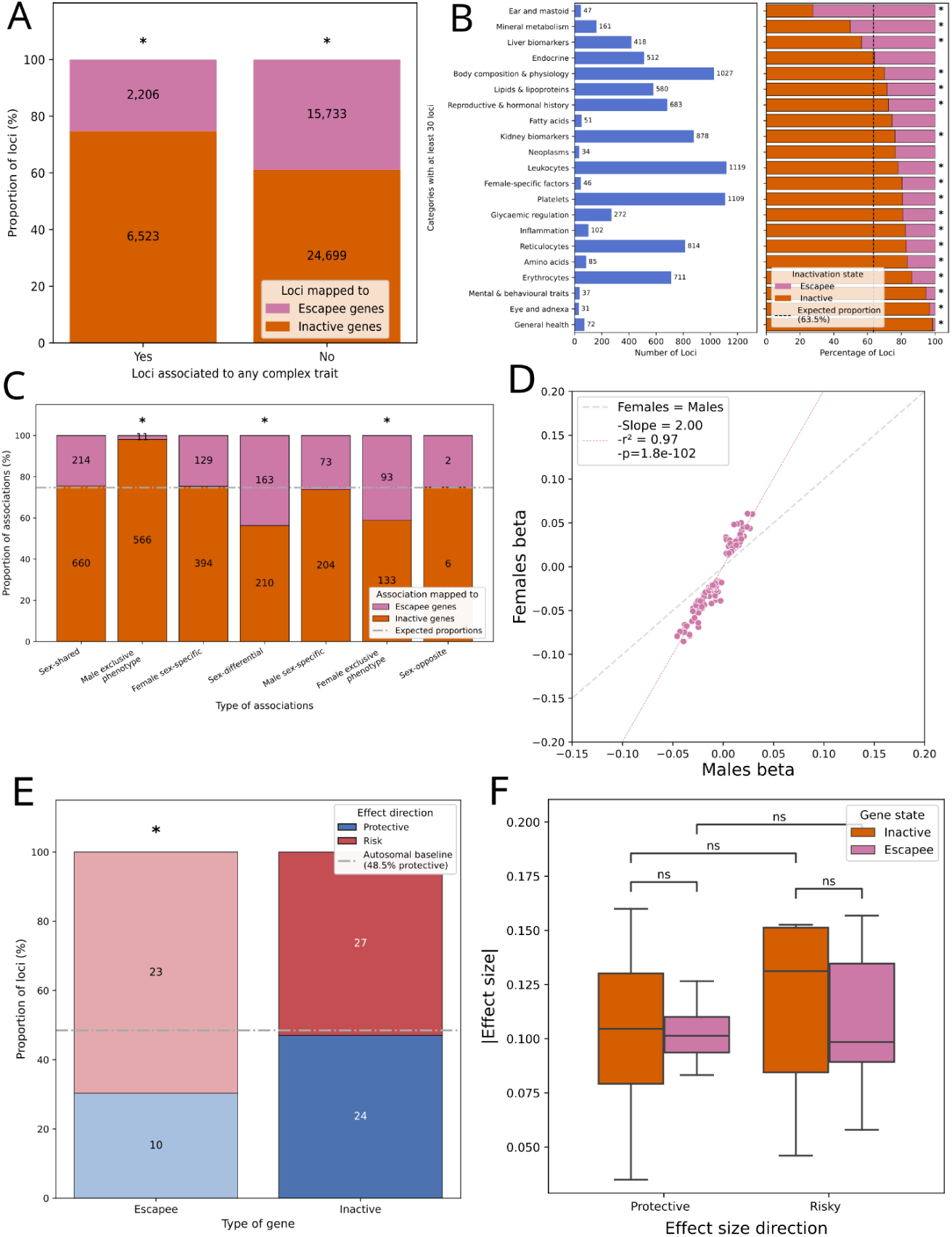
X-chromosome inactivation (XCI) status of trait-associated loci. Escape genes are shown in pink and inactive genes in orange; genes with uncertain XCI status were excluded. Asterisks (“*”) mark significant deviations from the expected proportion, while “ns”, not significant. **A)** Percentage of LD-pruned non-PAR Xchr loci mapped to escape and inactive genes among loci associated (left) and not associated (right) with any complex trait; numbers are loci counts. **B)** Left, number of LD-pruned non-PAR Xchr loci associated to phenotype category; categories with at least 30 loci are shown. Right, percentage of those loci mapped to inactive and escape genes; the dashed line marks the expected proportion of non-PAR Xchr loci mapped to inactive genes (63.5% inactive). **C)** Percentage of lead variants associations mapped to escape and inactive genes by sex-effect class and in male- and female-exclusive phenotypes; numbers are counts. The dashed line marks the expected escape proportion among associated loci (25%). **D)** Female versus male effect sizes of lead variants of sex-differential associations mapped to escape genes (n = 163). The pink line shows the linear regression (slope = 2, r² = 0.97, *p* = 1.8 × 10⁻¹⁰²) and the dashed grey line equal effects. **E)** Percentage of protective (β < 0, blue) and risk (β > 0, red) associations mapping to escape and inactive genes across 84 independent disease associations; numbers are counts. The dashed line marks the autosomal protective baseline (48.5%). **F)** Absolute effect sizes of disease associations mapped to inactive and escape genes, stratified by effect direction.

Functionally, trait-associated escapees and inactive genes were significantly enriched in 5 and 51 pathways, respectively, with no overlap between the two sets (Supplemental Table 15; Methods). Escapee-enriched pathways included “Estrogen metabolism” and “The activation of arylsulfatases”, whereas inactive-gene pathways included “Nucleotide metabolism” and “Ectoderm differentiation”. We next examined whether escapee loci were disproportionately concentrated in specific phenotype categories (Figure 5B; Supplemental Table 16). Escapee loci were present in 28 of 29 categories, but only three categories contained more escapee loci than expected (expected proportion = 36.5%), most notably “Ear and mastoid” (72%; two-proportion z-test *q* = 3×10⁻⁷) and “Mineral metabolism” (50%; two-proportion z-test *q* = 2×10⁻⁴). Conversely, 17 categories were enriched for inactive loci relative to expectation (63.5%), including “Eye and adnexa” (97%; two-proportion z-test *q* = 2×10⁻⁴) and “mental and behavioural traits” (94%; two-proportion z-test *q* = 1×10⁻⁴).

We next investigated the magnitude and direction of the genetic effects associated within escape and inactive genes. Overall, effect sizes were significantly larger at escapee-associated loci, with a 1.04-fold increase in median effect size relative to inactive genes (Wilcoxon rank-sum test p = 0.0003; Supplemental Figure 11). The expected impact of the biallelic expression of escape genes on effect sizes becomes more evident when we evaluate the distribution and enrichment of escape genes across associations with different patterns of sex-specificity (Supplemental Table 17). Whereas the proportion of escape genes among sex-shared associations closely matched expectations (observed = 24% vs expected = 25%; Figure 5C; two-proportion z-test *p* = 0.88), sex-differential loci showed a significant enrichment of escapee associations (observed = 44% vs expected = 25%; Figure 5C; two-proportion z-test *p* = 6×10⁻¹⁵), which showed effect sizes , on average, 2-fold larger in females than in males (Figure 5D; Pearson’s test *p* = 1×10⁻¹⁰²). None of these escapee associations displayed larger effect sizes in males than in females. Consistent with escapee genes largely being involved in female-specific biology, we observe that female-exclusive phenotypes were enriched for escapee genes (observed = 41% vs expected = 25%; Figure 5C; two-proportion z-test *p* = 1×10⁻⁷), in contrast to associations identified in male-exclusive phenotypes, which showed the opposite trend (2%; Figure 5C; two-proportion z-test *p* = 1×10⁻³⁶).

We next asked in which direction loci associated with escape genes influence female health. Classifying disease associations as risk (β > 0) or protective (β < 0), on the GWAS catalog, the autosomes accounted for 52% of risky and 48% protective associations. When comparing escapees and inactive with these background proportions, escapees are biased towards risky associations (70% risk vs 30% protective; two-proportion z-test *p* = 0.02), while inactive loci showed non-significant differences between the risky and protective categories, with 53% and 47%, respectively (two-proportion z-test *p* = 0.83; Figure 5E; Supplemental Table 18). GWAS Catalog showed the same pattern, with risk associations outnumbering protective ones for escapees genes in comparison with the autosomes (69% vs 31%; two-proportion z-test *p* = 0.002), but not for inactive genes (risk = 57% vs protect = 43%; two-proportion z-test *p* =0.25). When we stratified by effect direction, escapee and inactive associations showed comparable absolute effect sizes between protective and risky sets. (Figure 5F).

To identify potential sex biases towards the disease risk associated with gene inactivation status, we analysed disease associations showing different effects between sexes: the non-sex-shared category (sex-differential and female-specific together). Consistent with our findings, escape genes accounted for more risky associations than protective ones (risky = 64% vs protective = 36%). Unexpectedly, associations involving inactive genes showed the opposite pattern, with a strong predominance of protective over risk-increasing associations (protective = 89% vs risky = 11%; binomial test *p* = 0.0007; Supplemental Table 19).

### Chromosome X gene state prediction

Finally, to evaluate whether the differences in disease-association patterns between inactive and escape genes were sufficiently large to enable reliable discrimination between the two classes, we trained an XGBoost^39^ classification model using female-related association summary statistics, including association *p*-values, effect sizes, and standard errors, among others (Methods). 308 mapped genes to trait associations loci were retained for classification (220 for training and 88 for testing). Model performance was successful on the held-out test set, with 69 of 88 (78%) classified genes. The model correctly assigned 49 genes, including 45 inactive and 4 escape genes, corresponding to a weighted precision of 89% and weighted recall of 71% (Methods; Supplemental Figure 12). When applied to genes, for which the inactivation or escape status of the targeted gene remains unknown, we could classify a total of 618 lead variant associations targeting 158 genes, where 89 and 50 were labeled as inactive and escape, respectively (Supplemental Table 20).

Feature importance analysis indicated that the discrimination between escapee and inactivation status was driven predominantly by the sex-difference state of the signals and their inheritance model information (Figure 6B; Supplemental Table 21). Inactive patterns were mostly characterised by also showing association in males’ and by the recessive inheritance model, whereas escape genes were mostly associated with the additive and dominant ones (Figure 6C; Supplemental Figure 13). Consistent with the observed link between escape and non-recessive inheritance, loci classified under the dominant and the additive models were significantly enriched for escape loci with a 33% and 31%, respectively (expected = 25%; dominant two-proportion z-test *p* = 3×10⁻⁸; additive two-proportion z-test *p* = 1×10⁻¹¹; Figure 6D). These findings highlight the distinct sex-specific genetic signatures that differentiate escapee from inactive gene states.

**Figure 6.**
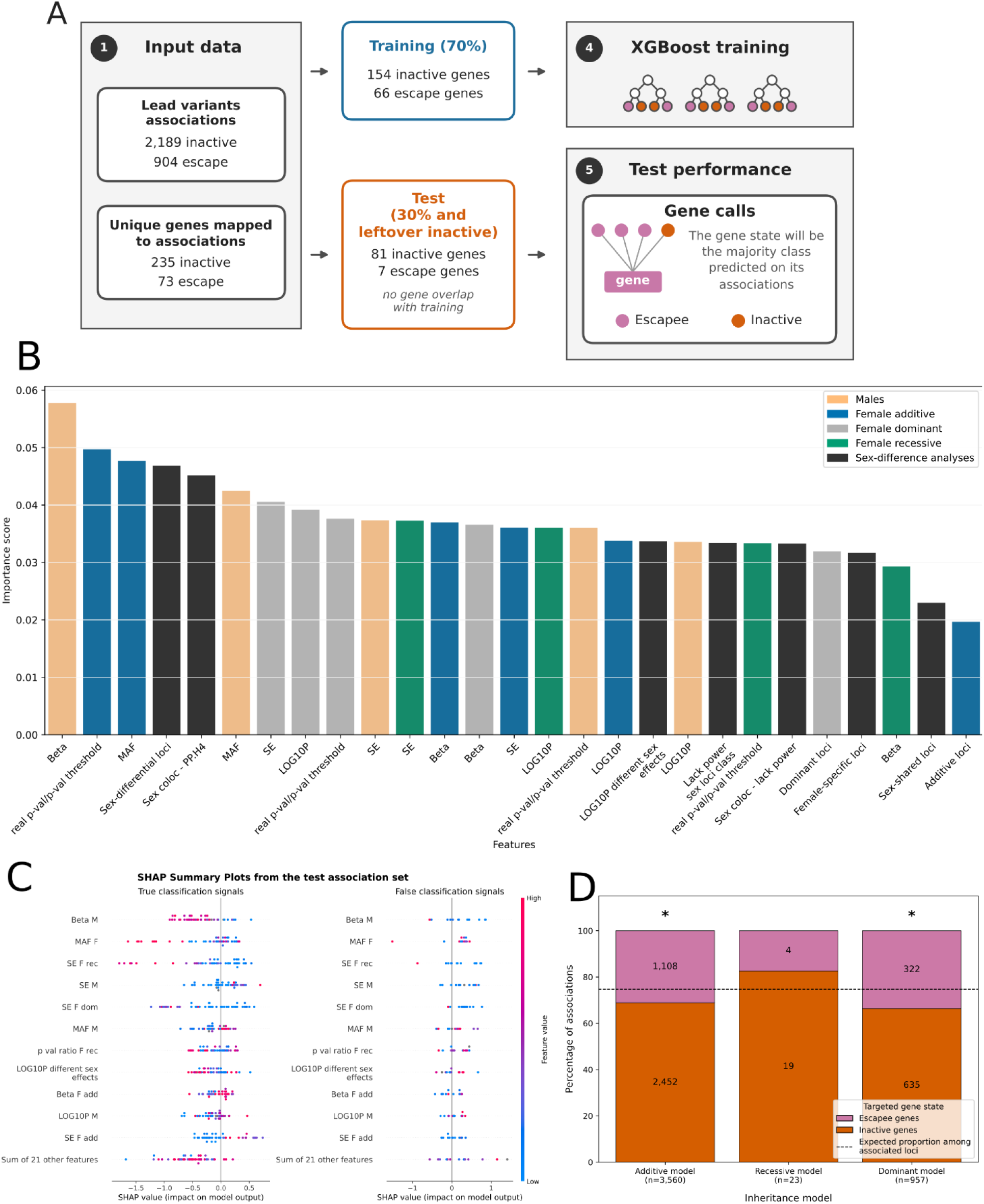
Prediction of XCI status from association patterns. **A)** Pipeline of the gene X-inactivation state classifier. Lead-variant associations mapped to genes of known state (inactive or constitutive escape) were split into training and test sets. The training process was done on an XGBoost model, and then, to evaluate performance on the test set, associations predictions were aggregated per gene by majority vote. **B)** XGBoost importance score of each feature, coloured by source: male PheXWAS (orange); female additive (blue), dominant (grey) and recessive (green) models; and sex-difference analyses (black). **C)** SHAP summary plots for test-set associations correctly (left) and incorrectly (right) classified. Each dot is an association, coloured by feature value (blue, low; pink, high); the 11 most important features are shown and the remaining 21 are summed. **D)** Percentage of associations mapped to escape (pink) and inactive (orange) genes by best inheritance model (additive, n = 3,560; recessive, n = 23; dominant, n = 957); numbers are counts. The dashed line marks the expected escape proportion among associated loci (25%); asterisks mark significant enrichment.

## Limitations

Several limitations should be considered when interpreting our findings. Although our study leverages one of the largest Xchr resources to date, sample size still limits the full characterization of its contribution to complex phenotypes, and larger cohorts will further improve signal resolution. Our analyses were restricted to non-ultrarare genotyped variants, and imputation constraints reduce the ability to capture rare and structural variation, which likely plays an important role in Xchr architecture. Current escape and inactive annotations enabled quantification of X–inactivation escape effects, yet a more complete and accurate classification will be essential to refine their contribution. Methodological constraints also limited escape–status prediction, leaving 67 genes unclassified; integrating complementary functional data may enhance future inference. Despite these limitations, this work represents the most comprehensive assessment of Xchr involvement in complex disease and highlights escape from X–inactivation as a widespread and potentially protective feature of Xchr biology.

## Discussion

In this study, we provide a population-scale characterization of the contribution of the non-PAR region of Xchr to complex traits by integrating sex-stratified genetic analyses across nearly 800,000 individuals and 3,867 complex phenotypes. In contrast with previous studies^3,4,7^, our results show that Xchr contributes to complex disease architecture to a comparable extent to that of autosomes of similar size. This suggests that its historical underrepresentation in GWAS mainly reflects systematic exclusion from studies rather than a reduced biological contribution.

Heritability measurements show an implication of Xchr across a broad range of disease categories, with sex-specific differences emerging for particular phenotypic domains, including nervous, circulatory, respiratory, and congenital diseases. Although theoretical models assuming shared causal effects between sexes predict higher heritability in males than in females^3,4^, our empirical analyses reveal a disease-dependent relation. In median, female heritability estimates exceed those in males, consistent with the presence of sex–differentiated effects that modulate the expected variance relationship. These observations suggest that deviations from the shared–effects model, driven by sex–specific genetic architecture, differential regulation or escape patterns, substantially shape Xchr heritability across complex traits.

We also identified thousands of independent X-linked associations across hundreds of traits, providing new genetic biomarkers and confirming the wide functional impact of this chromosome. The enrichment of dominant inheritance patterns among X-linked associations agrees with the XCI and the effective allelic composition in both sexes. Importantly, even though effect directions were generally aligned between sexes, sex-diferential loci associated with escape genes displayed stronger female effects, consistent with their increased dosage. Our analyses further reveal the relationship between XCI escape and complex trait susceptibility.

Escape genes were less frequently associated with disease loci than expected based on their genomic abundance, but when involved, they showed an enrichment for risky associations. These results suggest that escape from XCI contributes to increasing the risk of disease in females, which in turn could explain their underrepresentation among disease genes.

The distinct trait-association patterns between escape and inactive genes also enabled the development of a ML model to classify XCI status from genetic association features, allowing the annotation of previously unresolved X-linked genes. Previous studies have shown that long interspersed nuclear elements (LINEs), mammalian–wide interspersed repeats (MIRs), Alu elements, short ACG/CGT–containing motifs, and additional regulatory features enriched near transcription start sites can distinguish inactive from escape regions on Xchr^30,31^. Our study, based on complex–trait association patterns, extends and complements these observations. These results establish inactivation escape as an important factor of female-specific genetic architecture and encourage the incorporation of escape biology into future studies of complex disease, sex differences, and precision medicine.

## Methods

### UK Biobank cohort

UK Biobank is a large-scale biomedical database with 500,000 participants, most of them of European ancestry^40^. This study was performed based on the information from the last update of the cohort in 2023, and this cohort comprises individuals aged between 50-90 years which were recruited in diverse regions of the United Kingdom between 2006 and 2010. Study variables were derived from multiple sources, including hospital inpatient records, self-reported questionnaires, nurse-led interviews, clinical assays of biological samples, anthropometric measurements, and genotype array data.

From 2,431 available fields, 501 were manually selected and curated. Fields were categorized by UKBK as binary, categorical (single or multiple), or quantitative, and subsequently, we harmonized them into either disease outcomes (binary format) or disease-related clinical measurements (quantitative format). Categorical variables with an inherent ordinal structure were treated as quantitative measurements. Quantitative variables were transformed for the analysis by applying Rank Inverse Normal Transformation. Remaining categorical variables were either one-hot encoded when no control category was defined or binarized by contrasting each case category against a designated control. For each binary trait, we required a minimum of 100 cases in at least one of the two sexes. For traits belonging to a tree-structured class, controls were restricted to individuals without a diagnosis of the target disease or any other disease within the same branch. Finally, for binary traits, a minimum case-control ratio of 1:100 was maintained. When the ratio was lower, the oldest controls were selected until the desired ratio was achieved. This processing yielded 3,386 disease outcomes and 354 disease-related quantitative traits for downstream analyses.

### dbGaP cohorts

A total of 46 cohorts covering 36 different complex traits were initially selected and downloaded from the database of Genotypes and Phenotypes (dbGaP) under their respective project identifier (Supplemental Table 1). A trait was determined to be analyzable when, considering all cohorts for that trait, genotype and phenotype data were available for at least 10,000 individuals with at least 100 cases. For each cohort, an initial step of data cleaning and formatting was applied in order to obtain genotypic information in PLINK format^41^, keeping those individuals for whom both genetic and phenotypic information was available. As in UKBK, we maintain a minimum case-control ratio of 1:100, in case the ratio was lower, we selected the oldest controls until we achieve the desired ratio.

### Genetic quality control

A standard quality control was performed, both at the sample and at the variant level^16^. At the sample level, individuals were excluded based on the following filters: (i) genotype missingness ≥ 0.05; and (ii) ancestry outliers, which were identified using the 1000 Genomes Project reference panel^42^ and the smartPCA software^43^. At the variant level, variants fulfilling the following criteria were removed: (i) genotype missingness ≥ 0.05; (ii) minor allele frequency (MAF) ≤ 0.01; and (iii) Hardy-Weinberg disequilibrium (p<1e-20). A tailored quality control pipeline specific for the requirements of Xchr was implemented, consisting of filtering based on MAF ( ≤ 0,01 ) and genotype missingness (≥ 5%). These filters were performed on males and females separately. Then, variants in Hardy-Weinberg disequilibrium (p < 1e-20) were filtered out in non-affected female samples. Finally, variants that showed differences in frequency and genotype missingness between non-affected males and females (p < 1e-06) were excluded (Supplemental Table 2).

### Phasing and imputation

Xchr imputation across 47 cohorts was performed using GUIDANCE^15^, an integrated framework capable of conducting haplotype phasing and genotype imputation using one or multiple reference panels. First, the phasing step was performed with Shapeit(v2)^44^ in dbGaP and Shapeit(v4)^44^ in UKBB. Then, imputation was performed using Impute(v2)^45^. Importantly, due to the large size of the UKBB and the computational resources required for its analysis, data was split into chunks of 1,000,000bp. Both phasing and imputation were conducted using five reference panels; 1000G phase3^42^, GCAT^1^, HRC^46^, GoNL^47^ and UK10K^48^. The results obtained from the different panels were merged following the strategy in Alonso et al., 2021^33^, thus keeping the variants with the best info score obtained across all panels, and filtering the variants with MAF > 0.1%.

The resulting dataset for the association analysis consisted of genetic variants with an info imputation score ≥ 0.7 and MAF > 0.1% (Supplemental Table 2). This oxford format was converted into plink format files (hard-call-threshold = 0.9) using QCTOOLV2^49^.

### Association analyses

Sex-stratified genome-wide association analyses (GWAS) were performed separately for each complex phenotype using Firth logistic regression from REGENIE^34^. For association analyses of binary traits, the first 7 genetic ancestry principal components (PCs) were included as covariates. For quantitative disease-related measurements, age was included in addition to the same 7 PCs. In females, three inheritance models (additive, recessive, and dominant) were tested. To enhance statistical power, summary statistics from both sexes were subsequently analysed under the assumption of independence using a fixed-effects meta-analysis implemented in GWAMA^50^.

### *p*-value threshold estimation

N-of-1 permutation test thresholds were applied to PheXWAS and meta-analysis summary statistics from UK Biobank^36^. The 5% false discovery rate (FDR) thresholds were estimated after stratifying by sex, inheritance model (additive, recessive, dominant), phenotype type (binary or quantitative), and allele frequency (Supplemental Table 22).

For dbGaP cohorts, Bonferroni correction was applied. The number of independent loci was estimated after linkage disequilibrium pruning of imputed data using PLINK^41^ (r² = 0.8), accounting for both common and rare variants. Approximately 200,000 independent loci were obtained. To obtain a 95% confidence level, the significance threshold was calculated by dividing 0.05 by the number of independent loci. The resulting *p*-value threshold corresponded to 2.5×10⁻⁷.

### Heritability measures estimation

LD scores were computed with LD Score Regression(LDSC)^35^ following the Pan-UK Biobank guidelines^51^, stratifying by chromosome and sex. SNP-based heritability (h²) was estimated using LDSC^35^ on UKBK phenotypes with more than 5,000 individuals, using summary statistics from the additive model and restricting to genetic variants with MAF ≥ 0.01 outside the HLA region. Estimates were filtered using criteria consistent with Pan-UK Biobank, retaining h² values between 0 and 1 (accounting for standard errors) , λ_GC_ > 0.9, and ratio < 0.3. A Z-test was used to assess whether h² differed from zero, and *p*-values were adjusted using the Benjamini–Hochberg false discovery rate (FDR) method. Sex differences in non-sex-exclusive phenotypes were evaluated using a two-sample Z-test (Wald Z-test), with FDR correction applied to account for multiple testing.

### Chromosome Relative Involvement measure (CRI)

For each chromosome, phenotype and sex, significant heritability estimates (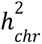) were adjusted for the number of imputed variants per chromosome (*N_chr_*). The corrected chromosome-specific h² values (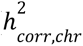) were then summed to obtain the total heritability (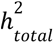) for each phenotype–sex combination. Finally, chromosome relative involvement (*CRI_chr_*) was defined as the percentage contribution of each chromosome’s corrected h² relative to the total h².

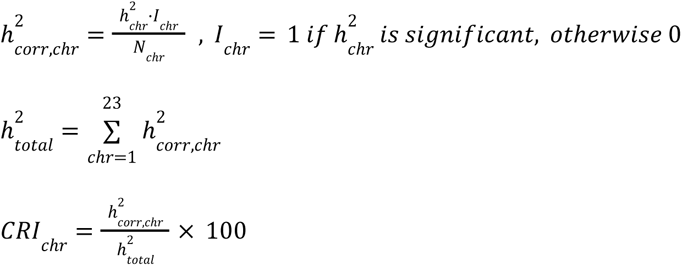

### Independent loci definition

For each phenotype, all genome-wide significant variants identified across the three GWAS analytical approaches (males, females, and meta-analysis) were aggregated.. These variants were subsequently clustered based on linkage disequilibrium using an r² threshold ≥ 0.8 with PLINK^41^. Each resulting linkage disequilibrium cluster was defined as an independent association signal and the variant with the lowest p-value was designated as the lead variant.

### Replication between UKBB and dbGaP cohorts

Phenotypes were manually harmonized between UK Biobank and dbGaP (Supplemental Table 7). Genome-wide significant loci identified in each cohort were tested for replication in the reciprocal cohort. Replication was defined as nominal significance (*p* < 0.05) for the same variant or for a proxy variant in linkage disequilibrium (r² ≥ 0.2).

### Sex-specific effect-size differences analysis

To assess the impact of sex on genetic associations, independent signals were classified into five categories, following the guidelines described in Khramtsova et al.^37^. First, colocalization analysis^52^ was performed to evaluate whether signals were shared between sexes, selecting the hypothesis with the highest posterior probability ( ≥ 0.7). Signals assigned to hypothesis 4 were considered shared (PP.H4 ≥ 0.7). Signals classified under hypothesis 1 or 2 were defined as male-specific (PP.H1 ≥ 0.7) or female-specific (PP.H2 ≥ 0.7), respectively.

Once an independent association was identified in both sexes, the equality of its lead variant effects was tested using a statistical test^53^. After correcting for multiple testing using the Benjamini-Hochberg method by type of phenotype (binary or quantitative), signals with equal effects were classified as sex-shared effect (q-value ≥ 0.3). Signals with different effects (q-value ≤ 0.05) were further divided into sex-differential or sex-opposite categories, depending on whether the effect direction was the same or opposite, respectively.

### Gene mapping

Colocalization analysis^52^ was performed between each independent GWAS signal and cis-eQTLs from GTEx v8^54,55^. Loci were defined as the signal’s variants ±1 Mb. Each signal was tested for every X-chromosome gene and GTEx tissue against the sex-stratified eQTL summary statistics (female, male, or sex-combined) matching the sex class of the GWAS signal. Sex-specific tissues were analysed with the corresponding single-sex data. Effect sizes, variances, allele frequencies, sample sizes, and trait standard deviations from both datasets were used. Signals with a posterior probability of a shared causal variant (PP.H4) > 0.7 were considered colocalized. In cases where multiple genes satisfied the colocalization criterion, the colocalized gene closest to the lead variant was retained as the target gene. If no gene met this threshold, the gene nearest to the lead variant.

### Gene annotation

Gene inactivation status was curated from a supplemental table compiling annotations from up to five independent studies^19,26,38,54,56^. For each gene, the consensus inactivation status was assigned by majority vote across the studies reporting on that gene. In cases of a tie, the gene was classified as “escape.” Genes classified as escape were further subdivided into “facultative escape” and “constitutive escape”: a gene was annotated as facultative escape if at least one of the contributing studies reported it as such, and as constitutive escape otherwise.

### Background escapees loci definition

All imputed genetic variants located on the non-pseudoautosomal region (non-PAR) of Xchr were pruned for linkage disequilibrium using PLINK^41^, with an r² threshold of 0.8 and a window size of 150 kb. A representative variant was thereby selected from each independent locus. Each representative variant was then annotated for linkage disequilibrium (r² ≥ 0.2) with any variant previously associated with a phenotype, using both our own association results and the GWAS Catalog^7^. Finally, loci were mapped to their closest gene through their selected variants; in cases where a gene was already implicated through colocalization in our association analyses, that gene assignment was added to that representative variant. Through this approach, each locus was classified into one of three categories: associated with our phenotypes, associated to GWAS Catalog phenotypes, or not associated with any phenotype. Moreover, each locus was annotated with the phenotype(s) with which it was associated with our XWAS analyses.

### Functional enrichment analyses

#### Pathways enrichment

All imputed genetic variants located on the non-pseudoFunctional enrichment analysis was performed on the set of discovered genes using g:Profiler^57^ with correcting by the default background and with Benjamini–Hochberg FDR correction for multiple testing. When genes were further stratified into subgroups (sex-specific categories or gene inactivation state), enrichment analysis was run separately for each subgroup. Enrichment was restricted to annotations from Gene Ontology (Biological Process and Molecular Function), Kyoto Encyclopedia of Genes and Genomes (KEGG), the Reactome Pathway Knowledgebase and WikiPathways.

#### Tissue expression enrichment

We download the TPM files of the GTEx (v11)^55^ and for each tissue, we computed the median TPM per gene, both sex-stratified and with sexes combined, and defined a gene as expressed in that tissue if its median TPM exceeded 5 and it was with at least 1 TMP in 80% of the samples. These tissue-specific gene sets were aggregated into a GMT file and supplied to g:Profiler^57^ as a custom annotation source. Enrichment analysis of the associated genes was then performed against this custom GMT file, with correcting by the default background and with Benjamini–Hochberg FDR correction for multiple testing.

#### Gene prediction model

The input dataset was prepared by removing significant associations that were not relevant to females. Specifically, associations classified as male-specific, those for male-exclusive phenotypes, and those lacking statistical power (female p-value ≥ 0.05) were excluded. Only signals located within 150 kb of the gene of interest, mapping to a single gene, and mapping to inactive or constitutive escapee genes were retained, resulting in 3,093 observations. For each association, the input consisted of the female PheXWAS summary statistics across all inheritance models, the corresponding male results, and the sex-differential analyses, together with one-hot encoded categorical features describing the sex-differential classification, the between-sex colocalization result, and the inheritance model. To reduce overfitting and mitigate class imbalance, a balanced set of associations was created and subsequently split into 70% training and 30% testing. Moreover, the rest of inactive associations excluded from the balancing step were added to the test set, but to prevent data leakage, associations mapped to genes present in the train set were excluded from the test set. The dataset sizes were 1,265 and 157 associations in the train and test sets, respectively. In addition, 631 associations targeted genes with unknown X-inactivation status.

An extreme gradient boosting (XGBoost)^39^ classifier was used to predict X-inactivation status. Its hyperparameter optimization was performed using a 5-fold cross-validation within a grid-search strategy over the following parameters: learning rate (0.01, 0.1, 0.2), maximum tree depth (2, 3, 4, 5), number of estimators (1-100), and subsample ratio (0.8, 1.0). The model selection was based on classification accuracy and the optimal hyperparameter configuration was: learning rate = 0.2, maximum depth = 4, number of estimators = 90, and subsample ratio = 0.8. The trained model was evaluated in the test set. Because multiple associations could map to the same gene, predictions were aggregated at the gene level with a majority-voting strategy. The X-inactivation state assigned to each gene corresponded to the class most frequently supported across associations.

The trained model was then applied to predict the X-inactivation status of all associations passing the filters described above. This enabled comparison of model predictions with existing literature for genes with a known state, and assignment of a new inactivation status to genes that previously lacked one (Supplemental Table 20).

Feature importance was initially assessed using the importance scores generated by the XGBoost model. Then, to further characterize feature contributions and improve model interpretability, SHapley Additive exPlanations^58^ values were calculated for all features. SHAP analyses were used to quantify the contribution of individual variables to model predictions at both the global and local levels, comparing feature contributions between correctly and incorrectly classified associations in the test set.

---

**Supplemental Figure 1.**
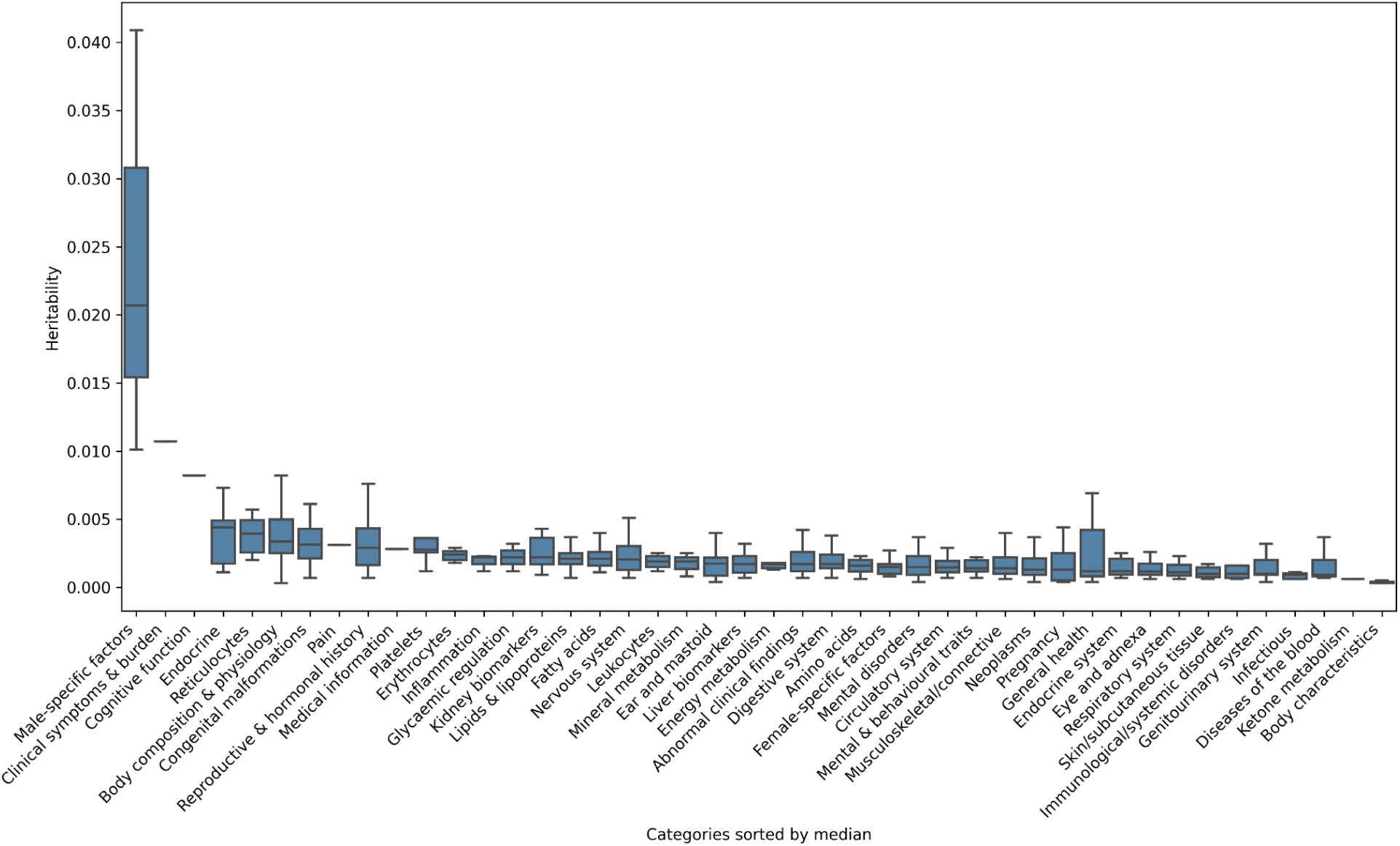
**Significant X-chromosome heritability across phenotype categories**. Significant h²_X_ for the 929 phenotypes in at least one sex, grouped by phenotype category and sorted by median.

**Supplemental Figure 2.**
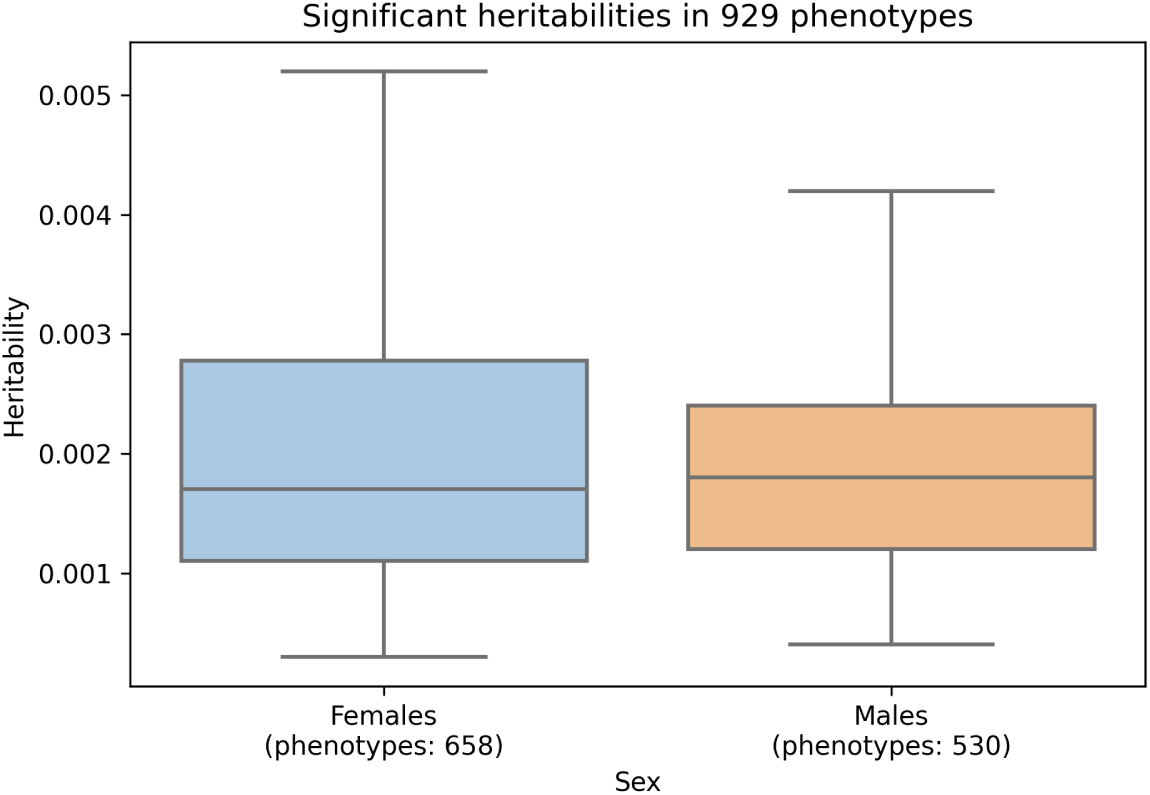
Sex comparison of X-chromosome heritabilities. Significant h²_X_ in females (blue; n = 658 phenotypes) and males (orange; n = 530) among the 929 phenotypes with significant h²_X_ in at least one sex (Mann–Whitney U p = 0.93).

**Supplemental Figure 3.**
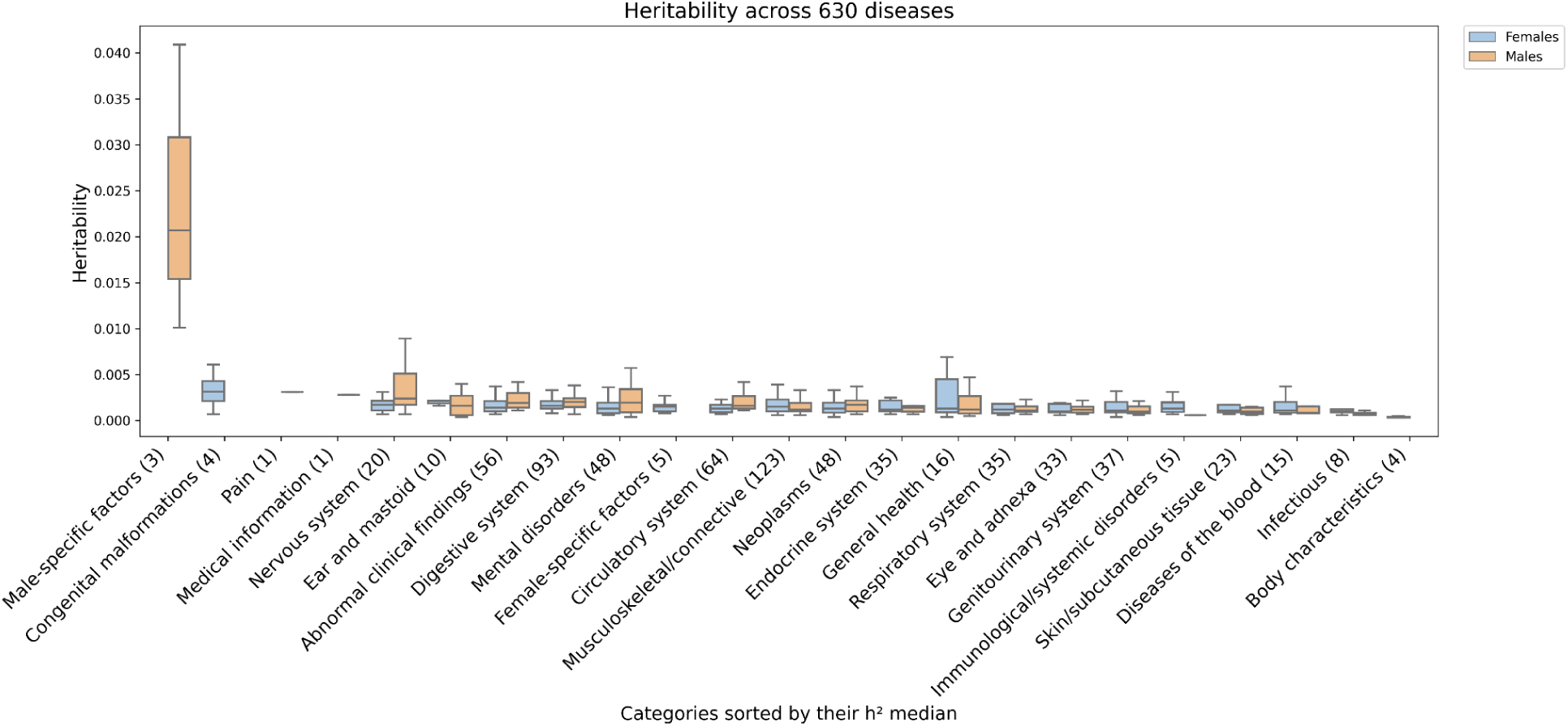
X-chromosome heritability across disease categories. Significant h²_X_ for 630 diseases grouped by category in females (blue) and males (orange), sorted by median; numbers in parentheses give the number of significant diseases per category of both sexes combined.

**Supplemental Figure 4.**
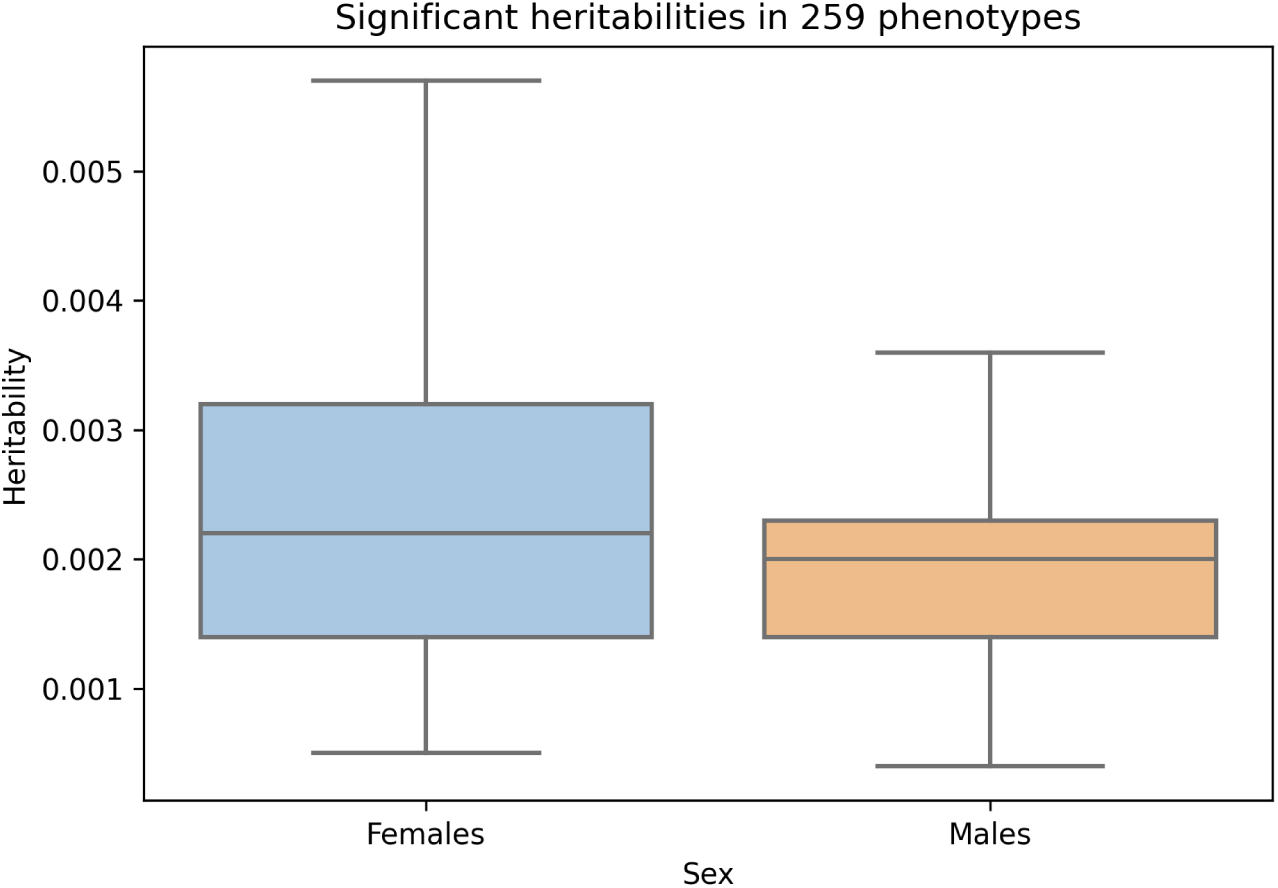
X-chromosome heritability in phenotypes with significance in both sexes. Significant h² in females (blue) and males (orange) for the 259 phenotypes with significant h²X in both sexes (paired t-test, corrected p = 4.4 × 10⁻¹¹).

**Supplemental Figure 5.**
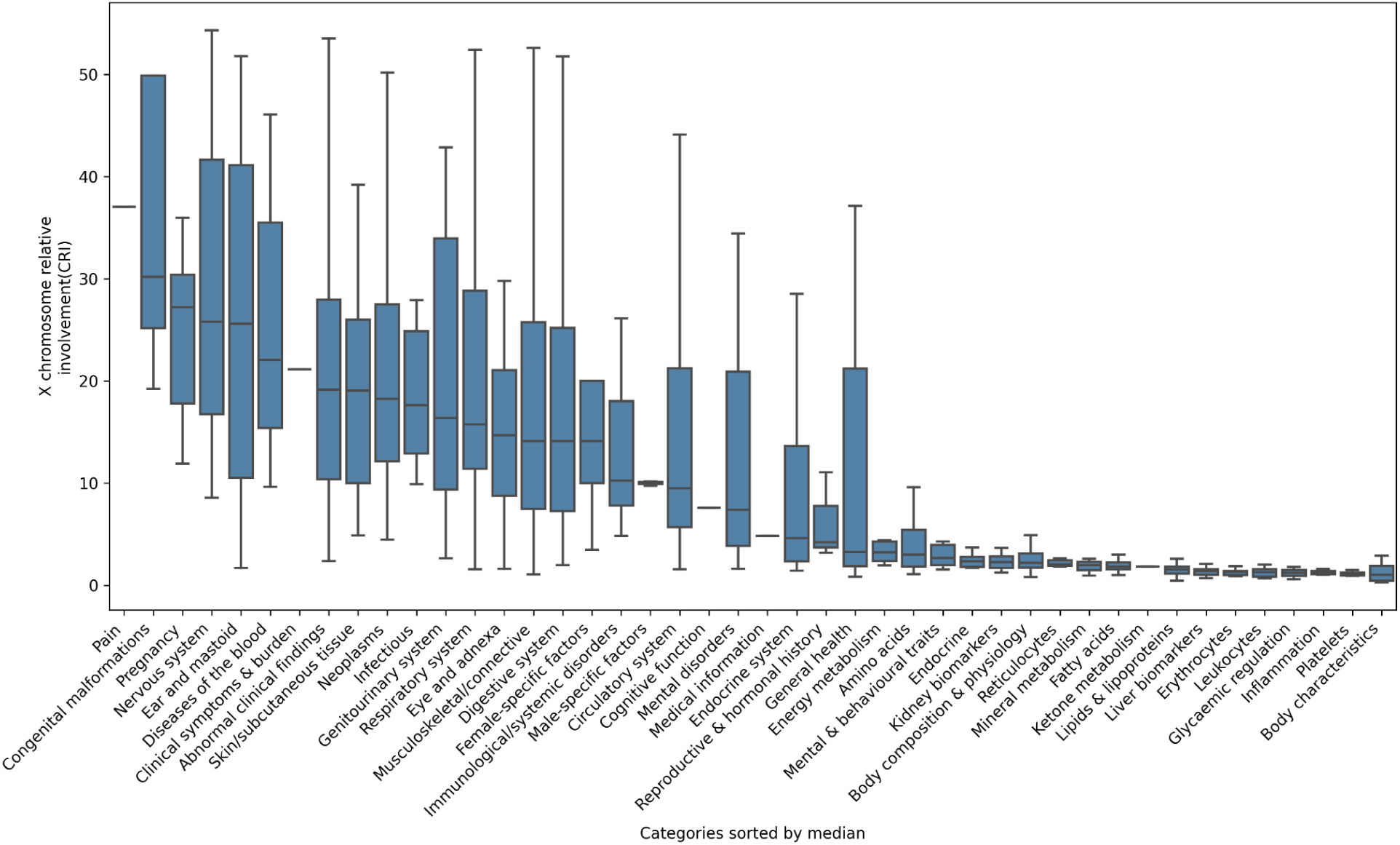
X chromosome relative involvement across 929 phenotype categories. Xchr CRI for the 929 phenotypes with significant h²_X_, grouped by phenotype category and sex; Sorted by median

**Supplemental Figure 6.**
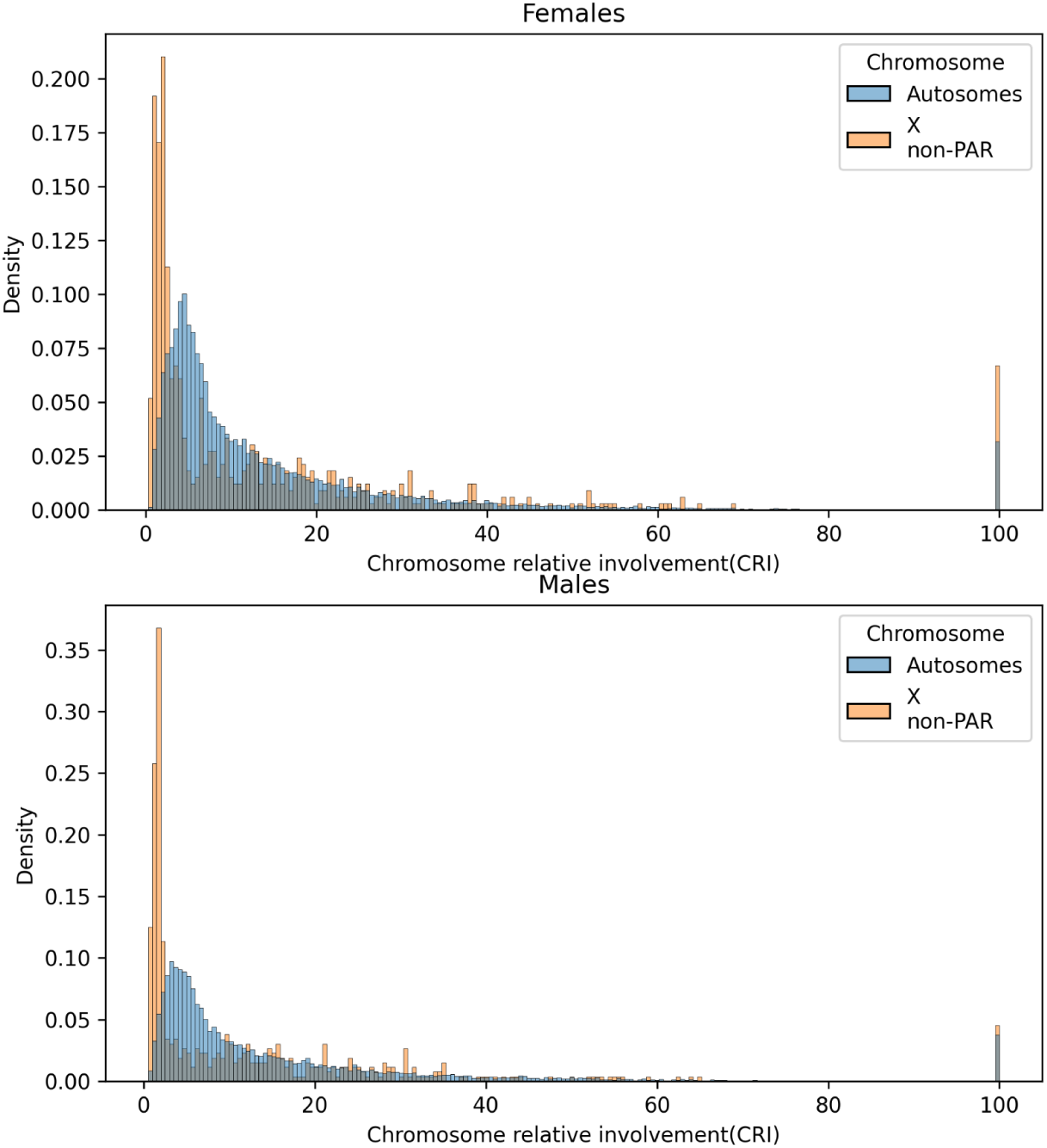
Comparison of chromosome relative involvement between X chromosome and autosomes by sex. Density of CRI for the Xchr (orange) and the autosomes (blue) in females (top) and males (bottom), in phenotypes with significant chromosomal heritability (median CRI_X_ = 5.3% vs CRI_A_ = 8%; Mann–Whitney U p = 2.4 × 10⁻³¹). The peak at 100% corresponds to phenotypes whose heritability is attributable to a single chromosome.

**Supplemental Figure 7.**
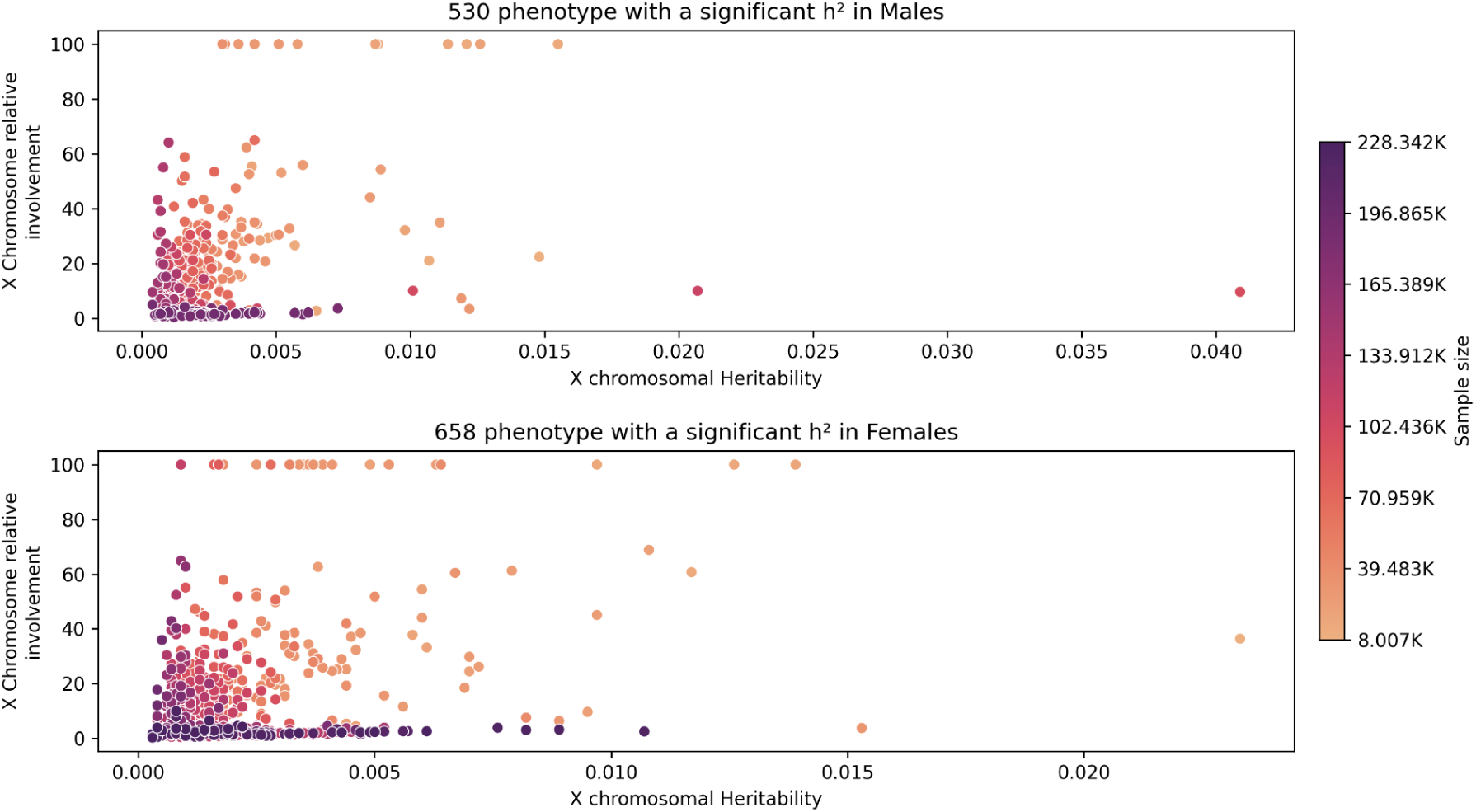
X-chromosome relative involvement by heritability. Xchr CRI against h²_X_ for phenotypes with significant h²_X_ in males (top; n = 530) and females (bottom; n = 658). Points are coloured by sample size.

**Supplemental Figure 8.**
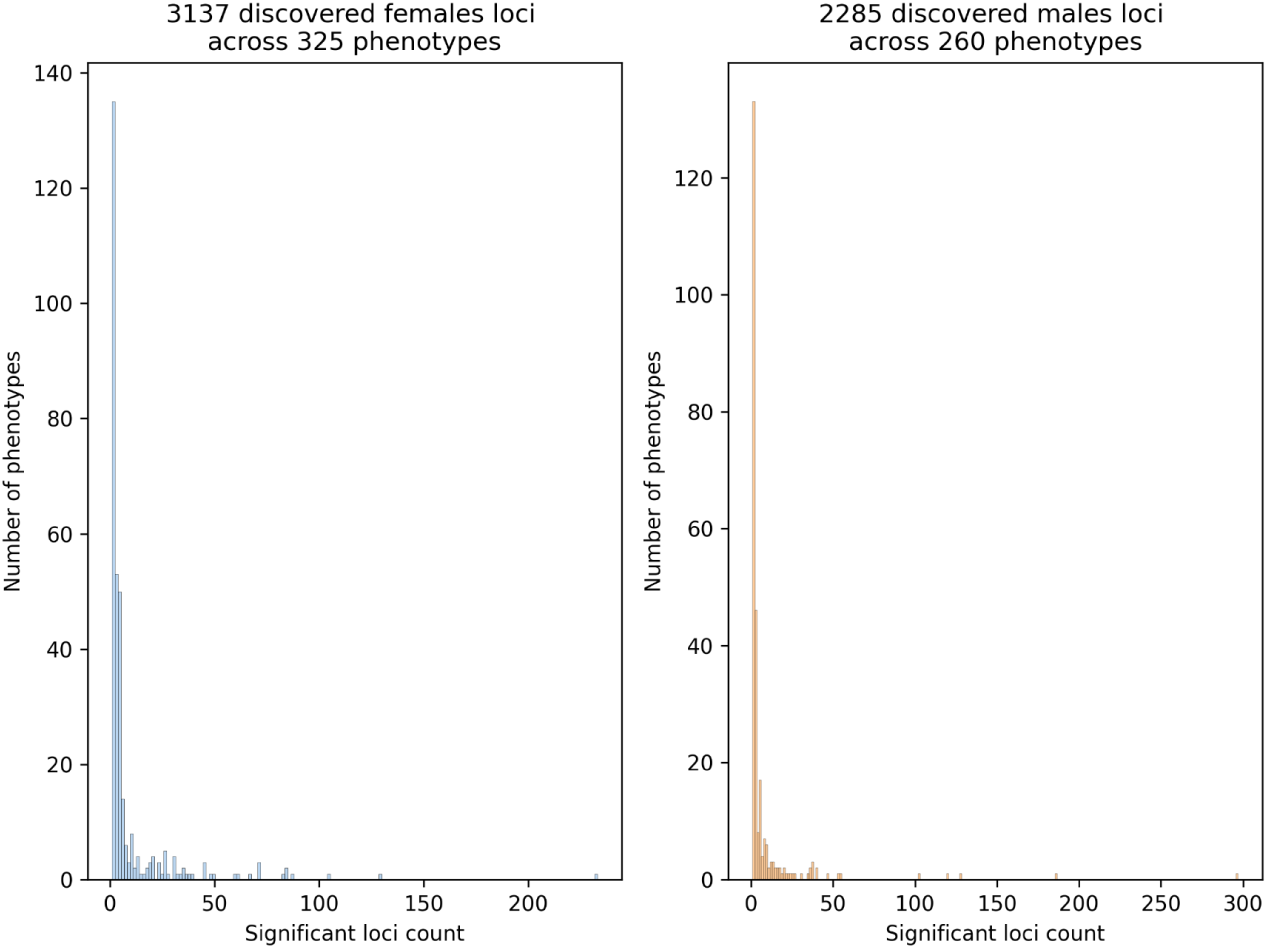
Number of significant loci per phenotype by sex. Distribution of the number of significant loci per phenotype for the 3,137 loci discovered in females (left; 325 phenotypes) and the 2,285 loci discovered in males (right; 260 phenotypes).

**Supplemental Figure 9.**
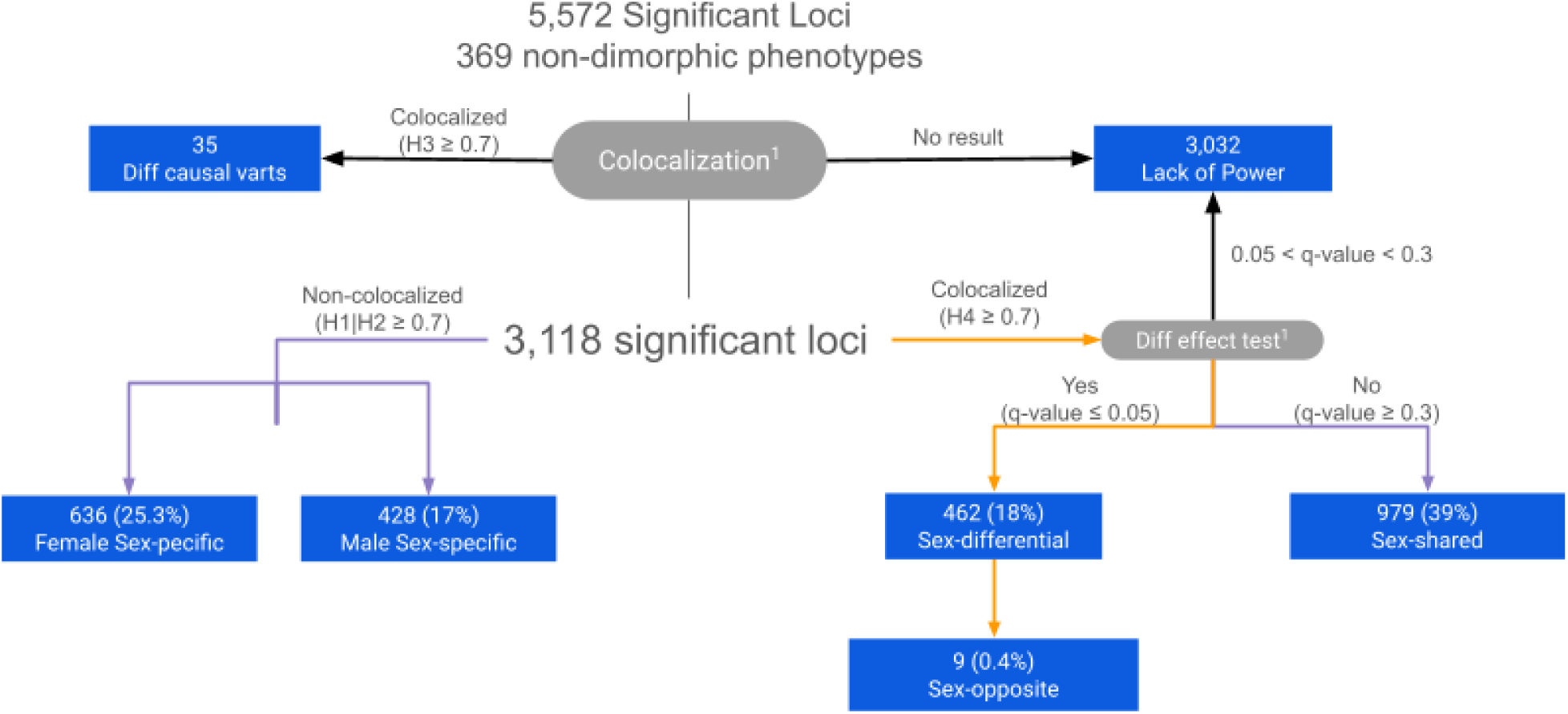
Classification of sex effects in non-sex-exclusive phenotypes. Pipeline applied to 5,572 significant loci in 369 non-sex-exclusive phenotypes. Female and male signals were first tested for colocalization (posterior probability ≥ 0.7). Loci without a colocalization result were classified as lacking power. 35 loci showed distinct causal variants in each sex (H3). Of the remaining 3,118 loci, those detected in one sex only were classified as female-specific (H2; n = 636) or male-specific (H1; n = 428). Loci shared between sexes (H4) were tested for effect-size differences and corrected by Benjamini–Hochberg q-value. If the q-value range between 0.05 and 0.3 in the effect-difference test, they were classified as lacking power. Otherwise, 979 were sex-shared (q ≥ 0.3), and 462 were sex-differential (q ≤ 0.05), including 9 with opposite effect directions (sex-opposite). Percentages are relative to the 2,505 loci in these final classes.

**Supplemental Figure 10.**
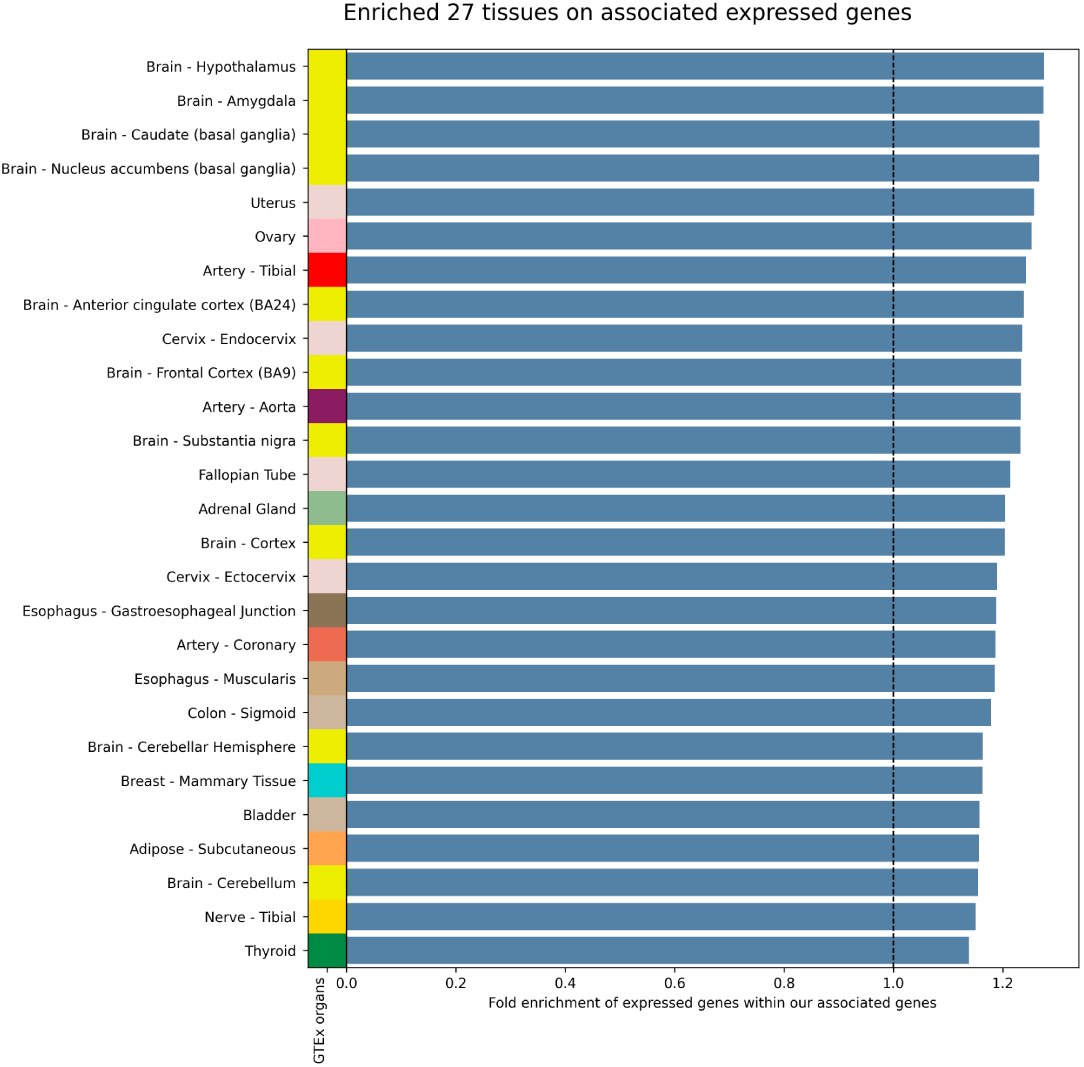
Tissue enrichment of X-chromosome associated complex-trait genes. Enrichment of X-chromosome genes mapped to complex-trait loci among the genes expressed in each GTEx (v11) tissue. Bars show the fold enrichment for the 27 tissues with significant enrichment (g:Profiler; Benjamini–Hochberg FDR < 0.05), and the coloured strip identifies each organ.

**Supplemental Figure 11.**
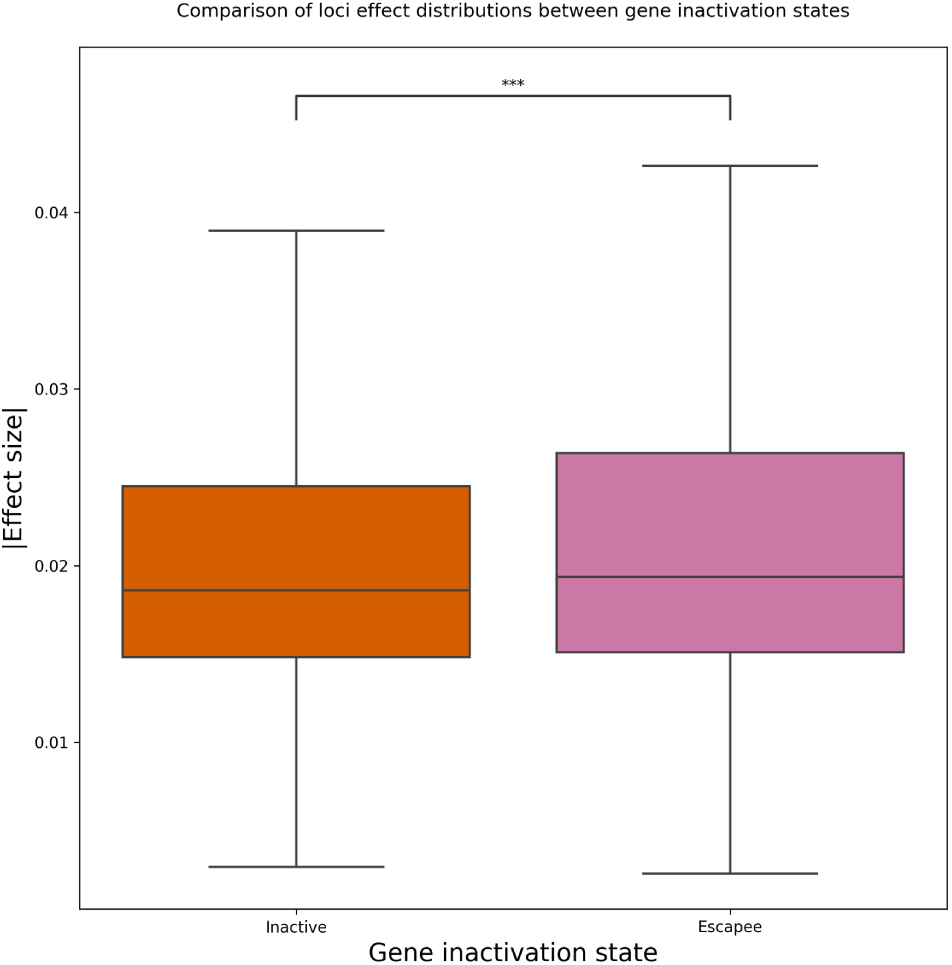
Effect sizes by XCI status. Absolute effect sizes (|β|) of loci mapping to inactive and escape genes. “***” Wilcoxon rank-sum test p = 0.0003.

**Supplemental Figure 12.**
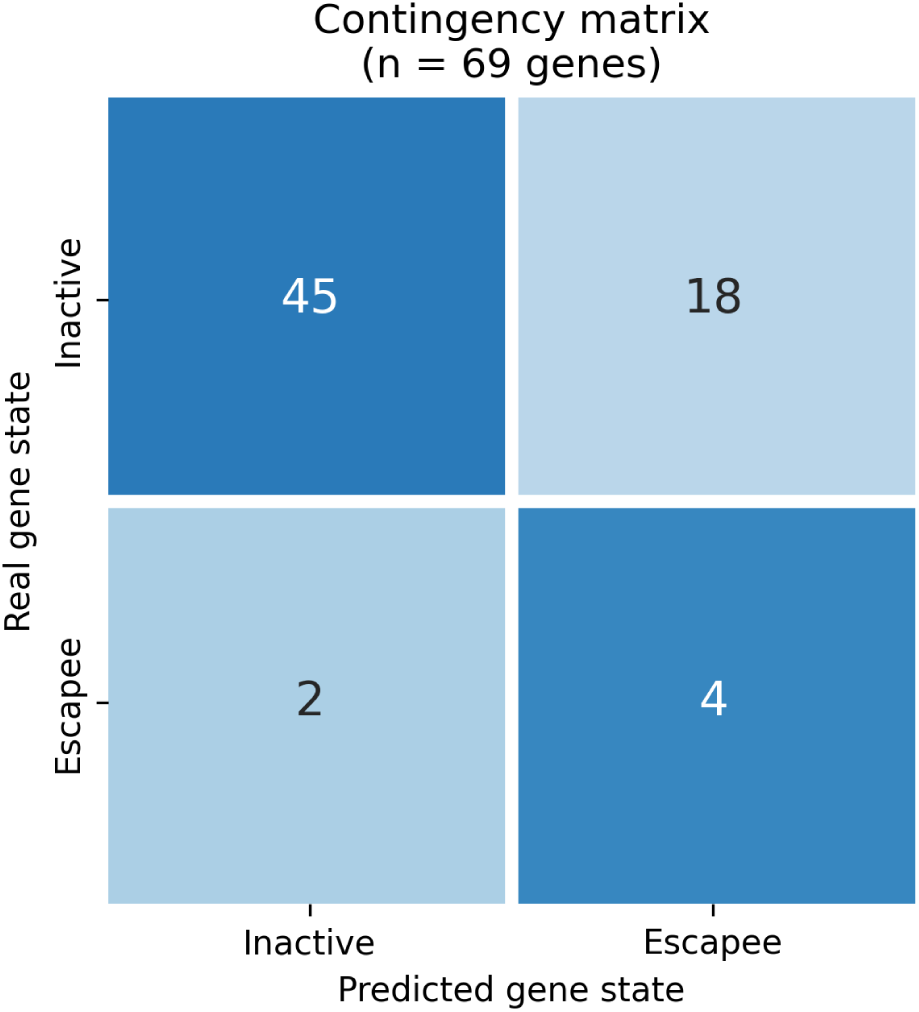
Performance of the XCI-status classifier in the test set. Confusion matrix of true versus predicted XCI status for the 69 test genes (of 88) that received a gene-level prediction. The model correctly classified 49 genes (45 inactive, 4 escape), with weighted precision 0.89 and weighted recall 0.71.

**Supplemental Figure 13.**
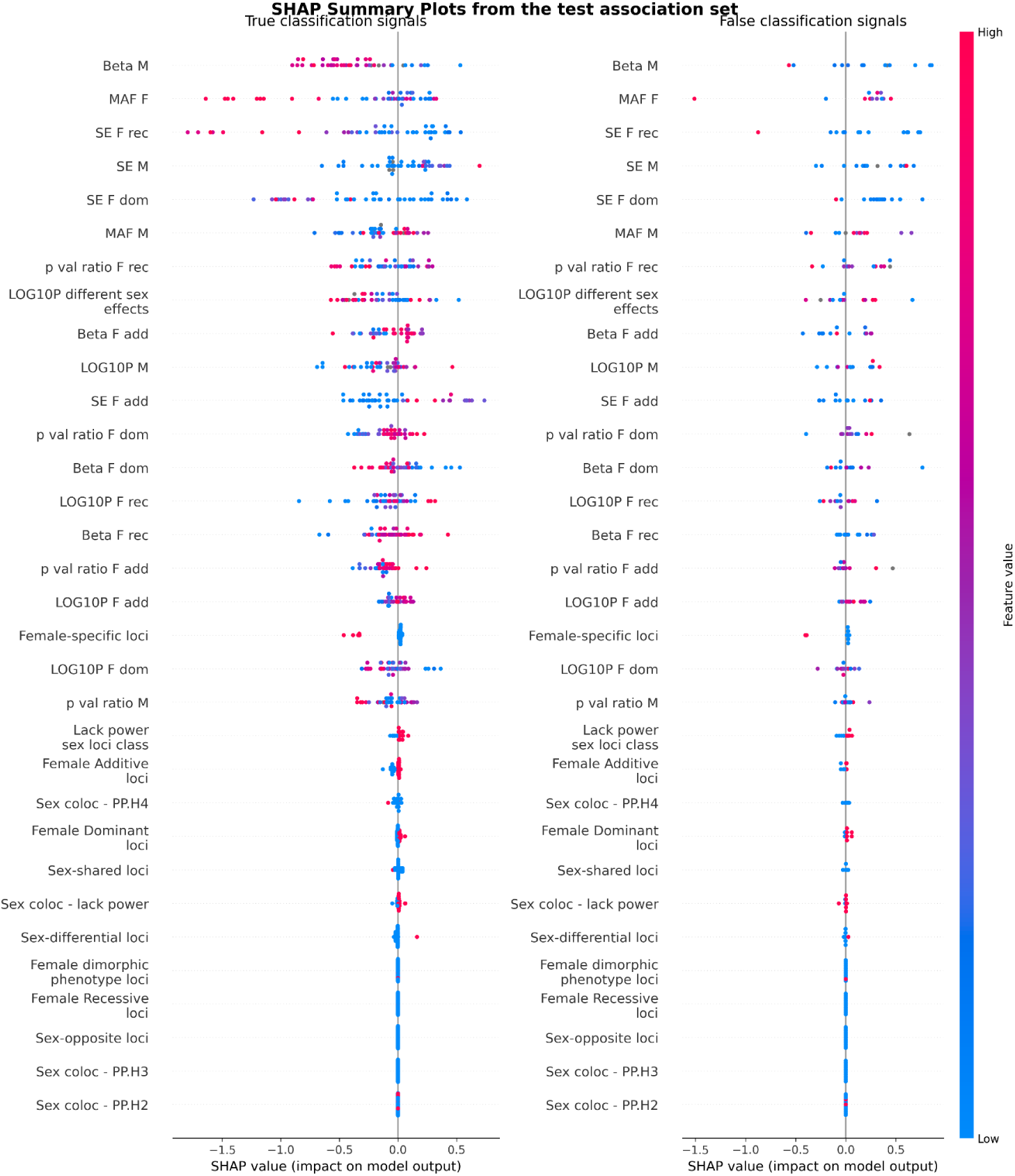
SHAP values for all model features. SHAP values of all features used by the XGBoost classifier, for test-set associations correctly (left) and incorrectly (right) classified. Each dot is an association, coloured by feature value (blue, low; pink, high).

## Data Availability

The complete summary statistics from this study have been deposited and are available to download at Zenodo and GWAS Catalog.

## Acknowledgments

We thank the participants and investigators of the UK Biobank study who made this work possible (Resource Application Numbers 1316623 and 85085). The Genotype-Tissue Expression (GTEx) Project was supported by the Common Fund of the Office of the Director of the National Institutes of Health, and by NCI, NHGRI, NHLBI, NIDA, NIMH, and NINDS. The data used for the analyses described in this manuscript were obtained from: v11 the GTEx Portal on 07/09/1998. The Genotype-Tissue Expression (GTEx) Project was supported by the Common Fund of the Office of the Director of the National Institutes of Health, and by NCI, NHGRI, NHLBI, NIDA, NIMH, and NINDS. The data used for the analyses described in this manuscript were obtained from: v11 the GTEx Portal on 07/09/1998.This work has been financed by the Science and Innovation Spanish Ministry under project PrevDis (PID2023-152867NB-I00). This work has been partially financed by the European Commission (EU-HORIZON NEARDATA GA.101092644). L.A-P fellowship within the “Generación D” initiative, Red.es, Ministerio para la Transformación Digital y de la Función Pública, for talent attraction (C005/24-ED CV1). Funded by the European Union NextGenerationEU funds, through PRTR. The technical support group from the Barcelona Supercomputing Center is gratefully acknowledged. Finally, we thank all the Computational Genomics group at the BSC for their helpful discussions and valuable comments on the manuscript.

## Contributions

**Conceived the work:** David Torrents, Miguel Ángel Pérez-Elena

**Designed the work:** David Torrents, Miguel Ángel Pérez-Elena

**Supervised the project:** David Torrents, Lorena Alonso-Parrilla, Miguel Ángel Pérez-Elena

**Acquired the data:** Miguel Ángel Pérez-Elena, Cecilia Salvoro, Daniel Matías-Sánchez, Lorena Alonso-Parrilla, Montserrat Puiggròs

**Prepared the data:** Miguel Ángel Pérez-Elena, Cecilia Salvoro, Daniel Matías-Sánchez

**Analysed the data:** Miguel Ángel Pérez-Elena, Leila Satari, Lorena Alonso-Parrilla, Cecilia Salvoro, Daniel Matías-Sánchez, Daniel Zugasti-Fernandez

**Interpreted the findings:** Miguel Ángel Pérez-Elena

**Supervised the findings:** Miguel Ángel Pérez-Elena, Lorena Alonso-Parrilla, David Torrents, Leila Satari

**Wrote the paper:** Miguel Ángel Pérez-Elena, Lorena Alonso-Parrilla, David Torrents

**Revised the paper:** All authors

## Notes

### Competing Interest Statement

The authors have declared no competing interest.

### Author Declarations

The study used (or will use) ONLY openly available human data that were originally located at: Uk biobank (Resource Application Numbers 1316623 and 85085) and dbGaP

